# Incidence of community acquired lower respiratory tract disease in Bristol, UK following the emergence of SARS-CoV-2: a prospective cohort study 2020-2024

**DOI:** 10.1101/2025.03.17.25324087

**Authors:** Anastasia Chatzilena, Catherine Hyams, Robert Challen, Elizabeth Begier, Jo Southern, Maria Lahuerta, Serena McGuinness, Madeleine Clout, James Campling, Jennifer Oliver, Andrew Vyse, Gillian Ellsbury, Prof Nick Maskell, Bradford Gessner, Adam Finn, Leon Danon

## Abstract

Surveillance of acute lower respiratory tract disease (aLRTD) is fundamental for understanding population health burden and healthcare needs. COVID-19 altered the epidemiology of respiratory infections, but post-pandemic aLRTD incidence and severity remain underexplored in the UK. We conducted a prospective cohort study of adults (≥18 years) admitted to two Bristol hospitals (August 2020–July 2024) with symptoms or a diagnosis of pneumonia, non-pneumonic lower respiratory tract infection (NP-LRTI), or no evidence of LRTI. Of 457,112 hospitalizations, 44,792 (9.8%) were due to aLRTD: 48.2% pneumonia, 35.2% NP-LRTI, and 16.7% no LRTI. Incidence peaked in 2021-22 (14.4/1,000 person-years) due to COVID-19 before stabilizing around 13.6. SARS-CoV-2 pneumonia declined; non-COVID pneumonia remained stable. Mortality risk was lower for NP-LRTI (HR 0.32) and no LRTI (HR 0.43) compared to pneumonia. Older age and comorbidities increased mortality. Non-COVID infections persisted despite interventions, emphasizing the need for surveillance and vaccination in public health planning.

## INTRODUCTION

Acute lower respiratory tract disease (aLRTD) is a major cause of morbidity and mortality globally, especially among older adults and those with underlying health conditions. The emergence of COVID-19 profoundly altered the landscape of respiratory disease^1,2^, influencing both the epidemiology and clinical presentation of aLRTD. Non-pharmaceutical interventions (NPIs) such as stay-at-home orders, facemask requirements, school and community closures and travel restrictions, along with widespread vaccination efforts, contributed to reduced transmission of many respiratory pathogens beyond SARS-CoV-2, including influenza and respiratory syncytial virus (RSV)^3,4^.

However, as these measures have been gradually relaxed, the post-pandemic dynamics of respiratory infections, including aLRTD, remain uncertain.

Published studies have examined the incidence of respiratory diseases during the COVID-19 pandemic, but few have evaluated the burden and severity of aLRTD as societies transition to an endemic phase of COVID-19. The impact of changes in viral circulation patterns, population immunity, and health system responses to respiratory disease has yet to be quantified. Understanding these trends is essential for guiding clinical and public health efforts, as changes in aLRTD incidence and severity can directly affect healthcare capacity and outcomes for affected populations.

Our previous analysis during the initial phase of the pandemic, highlighted the significant impact of SARS-CoV-2 on the incidence of community-acquired lower respiratory tract disease in Bristol, UK^5^. This manuscript provided valuable insights into the epidemiology of aLRTD during a period of intense public health interventions. Now, we aim to extend our understanding by examining the incidence and severity of aLRTD in the subsequent study years, as the UK transitioned from pandemic to endemic COVID-19. We conducted a prospective cohort study spanning August 2020 to July 2024 (during and after the pandemic), capturing aLRTD incidence among adults admitted to two acute care hospitals in Bristol, UK. Our primary objective was to determine accurate incidence of aLRTD(i.e., pneumonia, non-pneumonic lower respiratory tract infection (NP-LRTI) and cases with no evidence of LRTI), stratified by age group. We also quantified the contribution of COVID-19 to disease incidence, established non-SARS-CoV-2 aLRTD incidence rates, and evaluated changes in incidence during periods of shifting public health policies and viral circulation. Finally, we assessed the severity of these diseases to provide a comprehensive understanding of aLRTD during this unique epidemiological period.

## METHODS

### Ethics and permission

AvonCAP is a prospective observational cohort study of adults admitted to two large university hospitals in Bristol, UK. The study was approved by the Health Research Authority Research Ethics Committee East of England, Essex, reference 20/EE/0157, ISRCTN: 17354061.

Informed consent was obtained from cognisant patients, and declarations for participation from consultees for individuals lacking capacity. If it was not practical to approach individuals for consent, data were included under approval from the Clinical Advisory Group under section 251 of the 2006 NHS Act.

### Study design and study population

Adults (≥18 years) admitted to the two participating hospitals, which provide all acute secondary care in Bristol, between 1st Aug 2020 and 31st Jul 2024, were screened for inclusion as previously published^5^. Patients with signs and symptoms of respiratory disease, and those with three or more signs or a confirmed clinical or radiological aLRTD diagnosis, and disease ≤28 days in duration, were included. Relevant signs and symptoms were: documented fever (≥38°C) or hypothermia (<35·5°C); cough; increased sputum volume or discolouration; pleurisy; dyspnoea; tachypnoea; examination findings (e.g. crepitations); or, radiological changes suggestive of aLRTD in the opinion of a consultant radiologist, such as consolidation or pulmonary oedema. Patients were excluded from the study if their symptoms developed after 48 hours of admission or within 7 days of a previous discharge from hospital. Patients whose signs/symptoms were not attributable to aLRTD were excluded (e.g. fever and tachypnoea attributable to urosepsis). Eligible cases were classified as pneumonia, NP-LRTI or no evidence of LRTI.

Demographic and clinical data were collected from electronic and paper medical records and entered into an electronic clinical record using RedCAP^6^. Co-morbidity data, Charlson co-morbidity index^7^ (CCI; with published estimates of 10-year survival) and Rockwood clinical frailty score^8^ (with a score of 5–9 indicating frailty) were obtained, along with vaccination records from linked general practitioner (GP) records.

### Case definitions

Pneumonia was classified as acute respiratory illness with confirmed radiological changes compatible with infection or if the treating clinician made the diagnosis. In keeping with the National Institute for Health and Care Excellence (NICE) and British Thoracic Society (BTS) guidelines^9^ patients assigned a pneumonia diagnosis were counted as a pneumonia case even in the absence of radiological investigation or infiltrate on imaging, due to known false-negative radiology in pneumonia. NP-LRTI was defined as the presence of signs and symptoms of aLRTI in the absence of infective radiological changes and a clinical diagnosis of pneumonia. Under these case definitions, any patients with aLRTD signs or symptoms due to non-infectious aLRTD would have been assigned appropriately to chronic respiratory disease exacerbations (CRDE) or heart failure groups.

### Outcomes

We determined all-cause mortality within 30-days of hospital admission, hospital length of stay (days) and requirement for and length (days) of any intensive care or high-dependency unit (ICU/HDU) treatment. The population of Bristol was estimated as previously described, including full methodology^10^. Hospital admission data were linked to aggregated GP practice patient registration data within NHS Bristol, North Somerset and South Gloucestershire Clinical Commissioning Group for 2017–2019. The proportion of GP practices’ aLRTD hospitalisations that occurred at a specific study hospital was multiplied by their patient registration count for six age groups to obtain the practices’ contribution to that hospital’s denominator (e.g., if 50% of GP practice admissions were at a specific study hospital among persons 50−64 years, the practice contributed half of their patients 50–64 years to the denominator). The denominator was adjusted by the ratio of registered patients of each age group in 2019 versus each 3-month period during the study. Annual incidence was calculated per 1000 person years from 1st August 2020 to 31st July 2024, using the sum of case numbers (numerator) divided by the population (denominator) in each small time period. As the denominator calculation was performed for the subgroup of patients who resided in the Bristol Clinical Commissioning Group (CCG) area, patients from outside this area were excluded from the numerator for the incidence calculations.

### Statistical Analysis

The primary aim was to report incidence rates by clinical presentation in the period during and after the SARS-CoV-2 global pandemic, and to describe the new baseline for aLRTD post pandemic. Characteristics of all aLTRD admissions were described with categorical data presented as counts/percentages, and continuous data as either means/standard deviations (SD) or medians/interquartile (IQR) ranges, with normality of distributions determined with the Anderson-Darling test. aLRTD was stratified by clinical presentation; pneumonia, non-pneumonic lower respiratory tract infection (NP-LRTI) or no evidence of lower respiratory tract infection. These three categories may co-present with either heart failure or exacerbation of chronic respiratory disease, or both. Pneumonic and NP-LRTI groups may have evidence of SARS-CoV-2 as a causative pathogen, determined by PCR or lateral flow device.

All persons aged ≥18 years contributed to the denominator for incidence estimate calculations and are reported per 1000 person years for the whole cohort, and stratified by clinical presentation and SARS-CoV-2 status. Patients who declined consent were analysed as undifferentiated aLRTD. Estimates of admission incidence from weekly admission counts were made using maximum likelihood, assuming the observed case count was a Poisson distributed quantity with a time varying rate. The rate was estimated using a locally fitted order 2 polynomial using a logarithmic link function using the methods of Loader et al.^11^. Crude outcome rates for death, discharge, ICU admission and ICU length of stay are presented, stratified by clinical presentation. An adjusted survival analysis of death within 30 days and hospital discharge within 30 days (censored by death) was performed using a Cox proportional hazard model. All analyses were conducted using R^12^.

## RESULTS

### Study population

Among 457,112 hospitalizations during the 4-year study period, 44,792 admissions were attributed to aLRTD and we obtained data on 96% of participants, with only 4% declining consent. Of these, 20639 (48.2 %) presented as pneumonia, 15061 (35.2%) as NP-LRTI, and 7134 (16.7 %) had no evidence of LRTI (Figure 1). The cohort of patients hospitalised with all types of aLRTD primarily comprised elderly patients (median age 73 years, IQR 58-83), with 8.2% residing in care facilities (Table 1). Comorbidities were common; 45.2% had a Charlson Comorbidity Index (CCI) ≥4 and exacerbation of chronic respiratory disease was high (44.9 %). Additionally, 55.2 % were current or former smokers (Table 1). Patients with pneumonia were generally older (median age 74.5 years) than those with NP-LRTI (71 years). The NP-LRTI group had a higher proportion of patients aged 18-34 (11.3%) compared to the pneumonia group (5.1%). Care home residency was more common in the pneumonia (10%) and NP-LRTI (7.3%) groups. Heart failure was most prevalent in the evidence of LRTI group (44.2%), as was exacerbation of chronic respiratory disease (49.9%) compared to the pneumonia group (42.2%). For patients’ characteristics by presentation see Supplementary Data S1-S4.

**Table 1:**
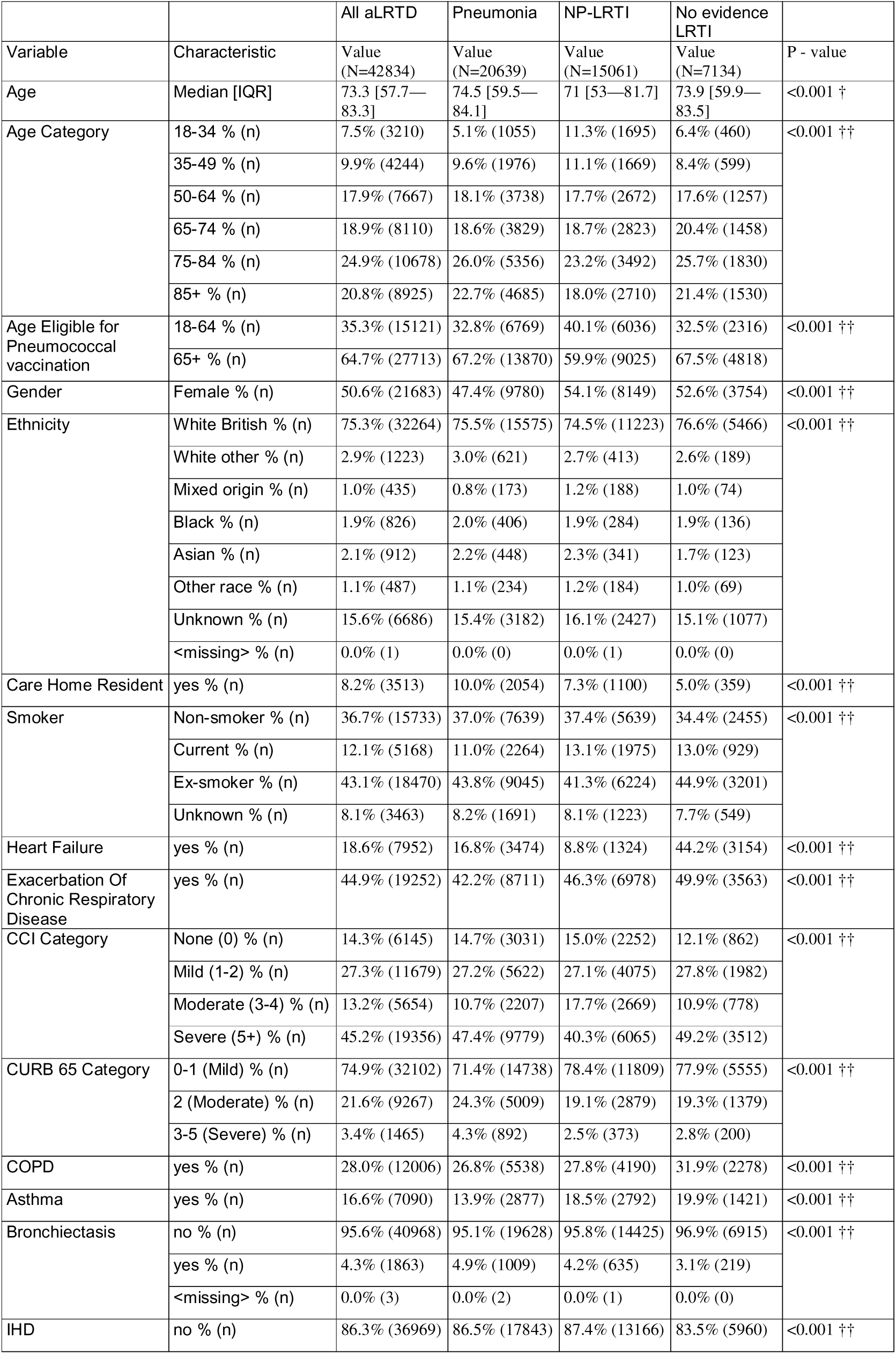
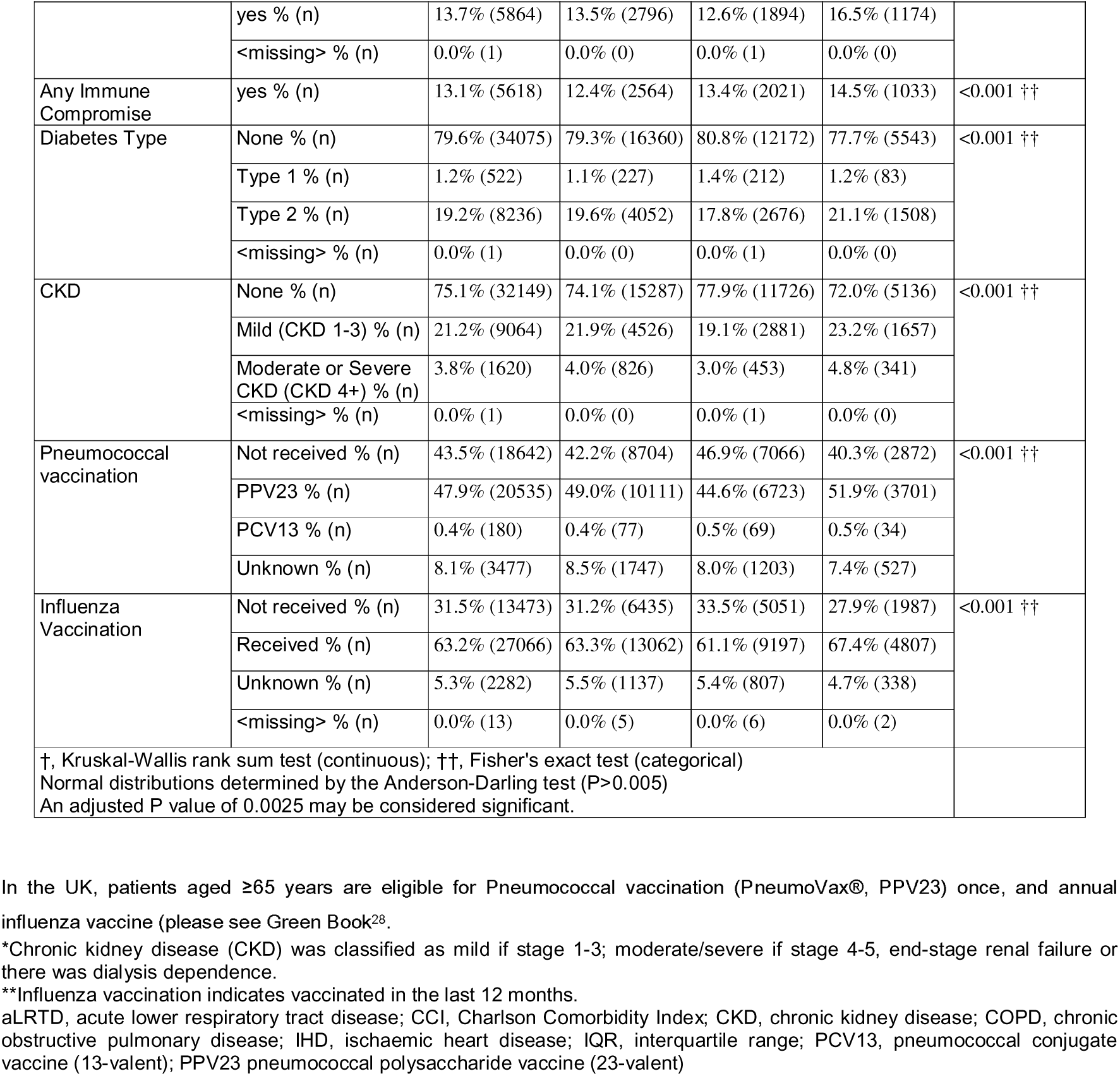
Characteristics of patients hospitalised with aLRTD for the period Aug 2020 to Jul 24.

**Figure one:**
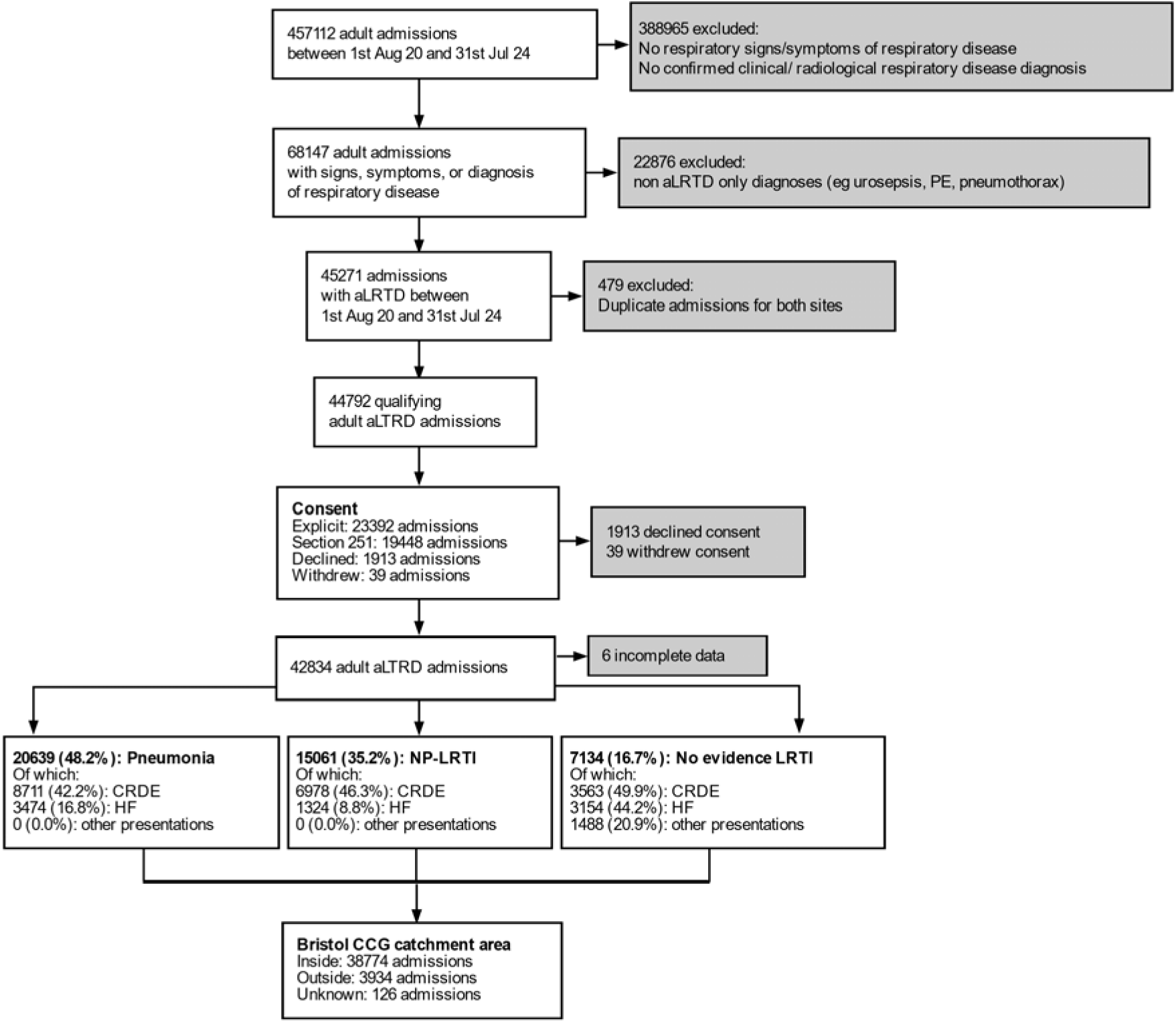
Study Flow Diagram. aLRTD, acute lower respiratory tract disease; CCG: Clinical Commissioning Group; CRDE, chronic respiratory disease exacerbation; HF, heart failure; LRTI, lower respiratory tract infection; NP-LRTI, non-pneumonic lower respiratory tract infection; PE, pulmonary embolus

### Incidence by age and study year

Over the four-year study period, the annual incidence rates for aLRTD among adults remained largely stable, with fluctuations primarily driven by COVID-19 cases (Table 2). The incidence of aLRTD peaked in Year 2 (2021-22) at 14.5 per 1,000 person years, up from 12.3 in Year 1, before stabilizing around 13.4 in subsequent years. Overall pneumonia incidence rates remained constant, although, amongst pneumonia cases, confirmed SARS-CoV-2 pneumonia exhibited the most substantial change, with an incidence rate of 2.5 per 1,000 in Year 1 that decreased to 0.9 by Year 4. Pneumonia with no evidence of SARS-CoV-2 fluctuated, ranging between a low of 3.6 in 2021-22 to 5.5 in 2023-24. NP-LRTI incidence increased in Year 2, largely due to COVID-19-related cases. Confirmed SARS-CoV-2-associated NP-LRTI rose, while NP-LRTI with no evidence of SARS-CoV-2 remained stable around 2.8 to 3.3. Cases with no evidence of lower respiratory tract infection showed minor variation, maintaining incidence rates around 2.1 to 2.4 per 1,000. Incidence rates per 100,000 per year are in Supplementary Data S5.

**Table 2:**
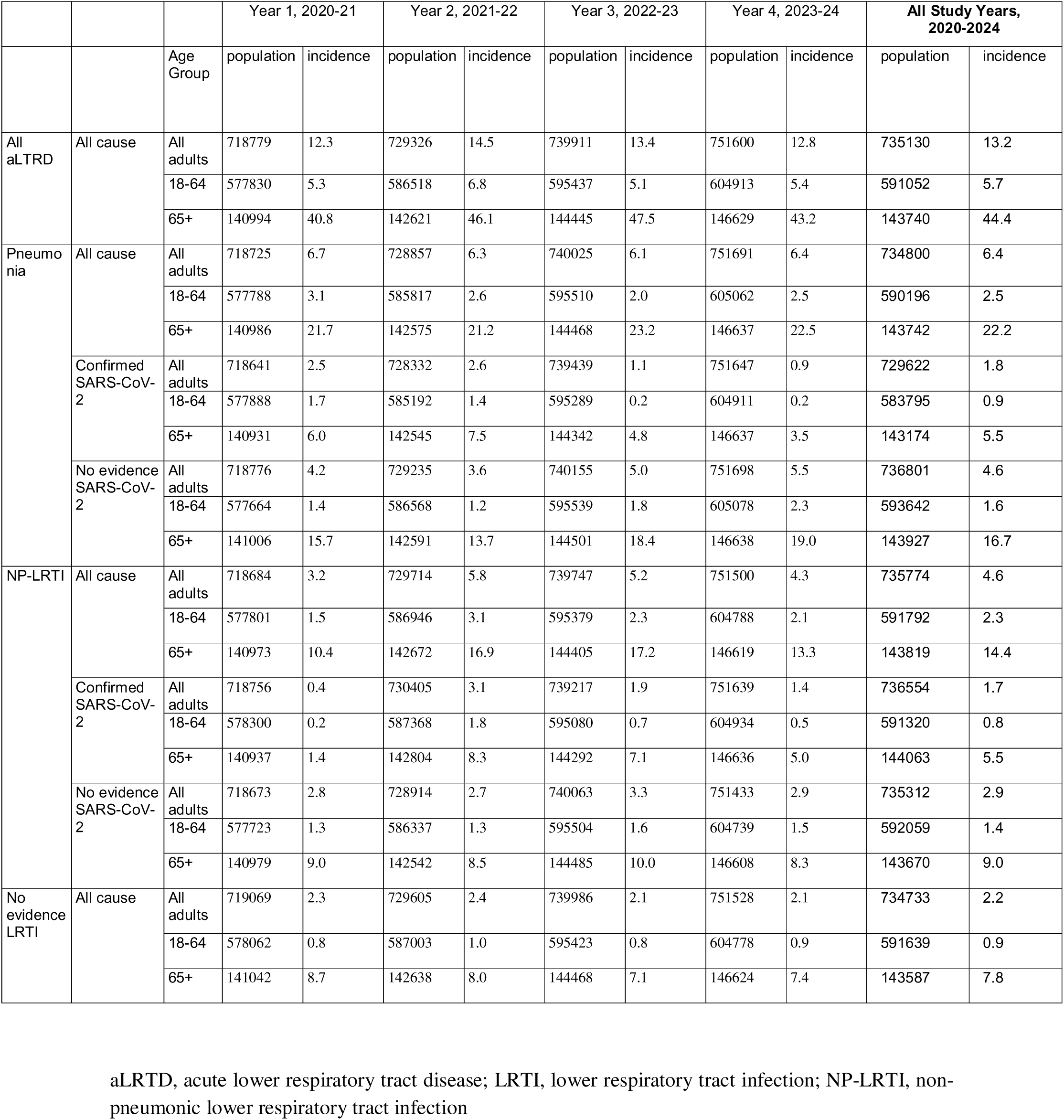
Overall incidence of aLTRD for the period Aug 2020 to Jan 2024. The average incidence of aLRTD per study year, stratified by clinical presentation (pneumonia, non-pneumonic lower respiratory tract infection (NP-LRTI), no evidence of lower respiratory tract infection (LRTI)), causative aetiology (all cause, confirmed SARS-CoV-2, no evidence of SARS-CoV-2) and age categories, and expressed as cases per 1000 person years.

Figure 2 shows the weekly incidence of aLRTD per 1,000 person-years, categorized into pneumonia, NP-LRTI, and no evidence of LRTI, stratified by age group and SARS-CoV-2 status. Incidence rates of all aLRTD subgroups fluctuated over time, with peaks aligning with the first pandemic waves in the UK during 2020–2022, as well as additional peaks reflecting seasonal trends in other respiratory infections (Figure 2A). Case rates were highest in the older age groups (65-74, 75-84, and 85+), especially during periods of high community transmission. Across all age groups, pneumonia exhibited the highest peaks in incidence, followed by NP-LRTI, while cases with no evidence of LRTI remained more stable.

**Figure 2:**
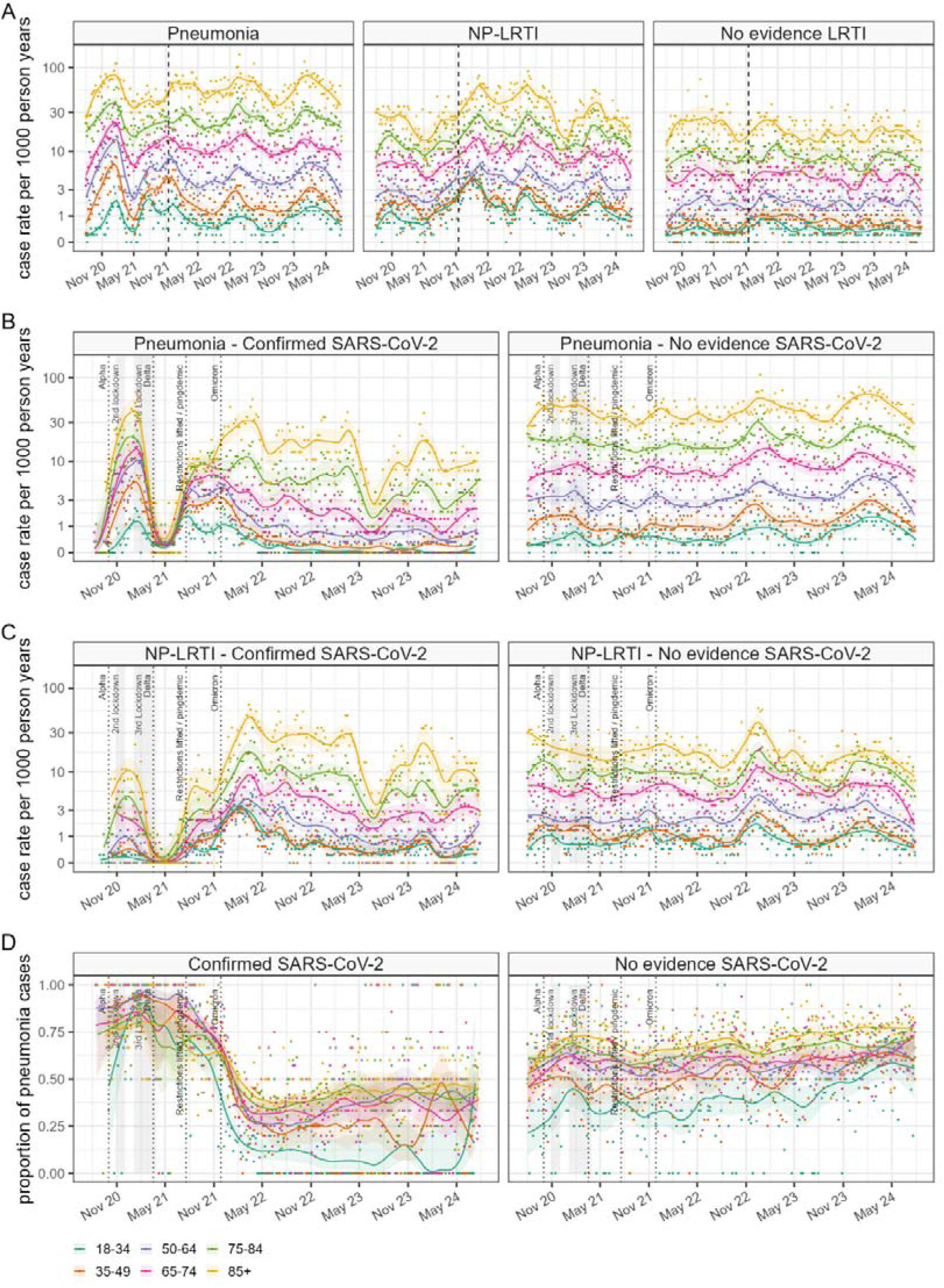
Weekly case rates of aLRTD Hospitalisations between Aug 2020 and Jul 2024 by Age Group. (A) Weekly cases rates of aLRTD stratified by clinical presentation for pneumonia, non-pneumonic lower respiratory tract infection (NP-LRTI) and cases with no evidence of lower respiratory tract infection (LRTI). Rates are stratified by age category and expressed as new cases per week per 1,000 person-years. The dashed line indicates data already published^5^. (B) Weekly cases rates of pneumonia stratified by SARS-CoV-2 PCR status. (C) Weekly cases rates of NP-LRTI stratified by SARS-CoV-2 PCR status. (D) Proportion of pneumonia cases stratified by SARS-CoV-2 PCR status.

Among pneumonia cases with confirmed SARS-CoV-2, incidence rates showed substantial increases during the first pandemic waves (Figure 2B). Increased pneumonia cases, particularly in older adults (especially those 85+) in late 2020 were followed by a decrease as public health restrictions were introduced. Pneumonia with no evidence of SARS-CoV-2 showed more consistent incidence rates, with slight increases in the winter seasons. NP-LRTI cases associated with SARS-CoV-2 followed similar trends, with peaks aligning with COVID-19 waves, though the severity was generally lower (Figure 2C). NP-LRTI incidence rates reduced during the second half of 2020 and into 2021. After 2022, NP-LRTI cases without evidence of SARS-CoV-2 were comparatively stable across age groups, although wintertime seasonal increases continued to occur. During early COVID-19 waves there was an elevated proportion of pneumonia cases among confirmed SARS-CoV-2 infections while a lower proportion was seen in later waves, such as those driven by the Omicron variant (Figure 2D). The proportion of pneumonia among cases with no evidence of SARS-CoV-2 remained relatively stable, with a slight upward trend over time.

### Outcomes

Outcomes were categorized by age groups (18-64 and 65+ years) (Figure 3). The 65+ age group consistently experienced longer hospital stays compared to the 18-64 age group (Figure 3A). The length of stay remained relatively stable both during the pandemic and in subsequent years. The 65+ age group had longer ICU stays, with apparent peaks (Figure 3B). There were noticeable spikes in ICU admissions, reflecting surges in severe cases of aLTRD, with a higher percentage of such admissions observed in the 18-64 age group than the 65+ age group (Figure 3C). Despite fluctuations, the overall rate of ICU admissions remained stable during the pandemic and in the following years. Regarding mortality outcomes, the 65+ age group demonstrated higher mortality rates throughout the study period, with specific intervals of increased rates appearing to align with peaks in ICU admissions and duration. Mortality remained relatively stable during and after the pandemic (Figure 3D-E). Detailed results for each study year are in Supplementary Data S6-S10.

**Figure 3:**
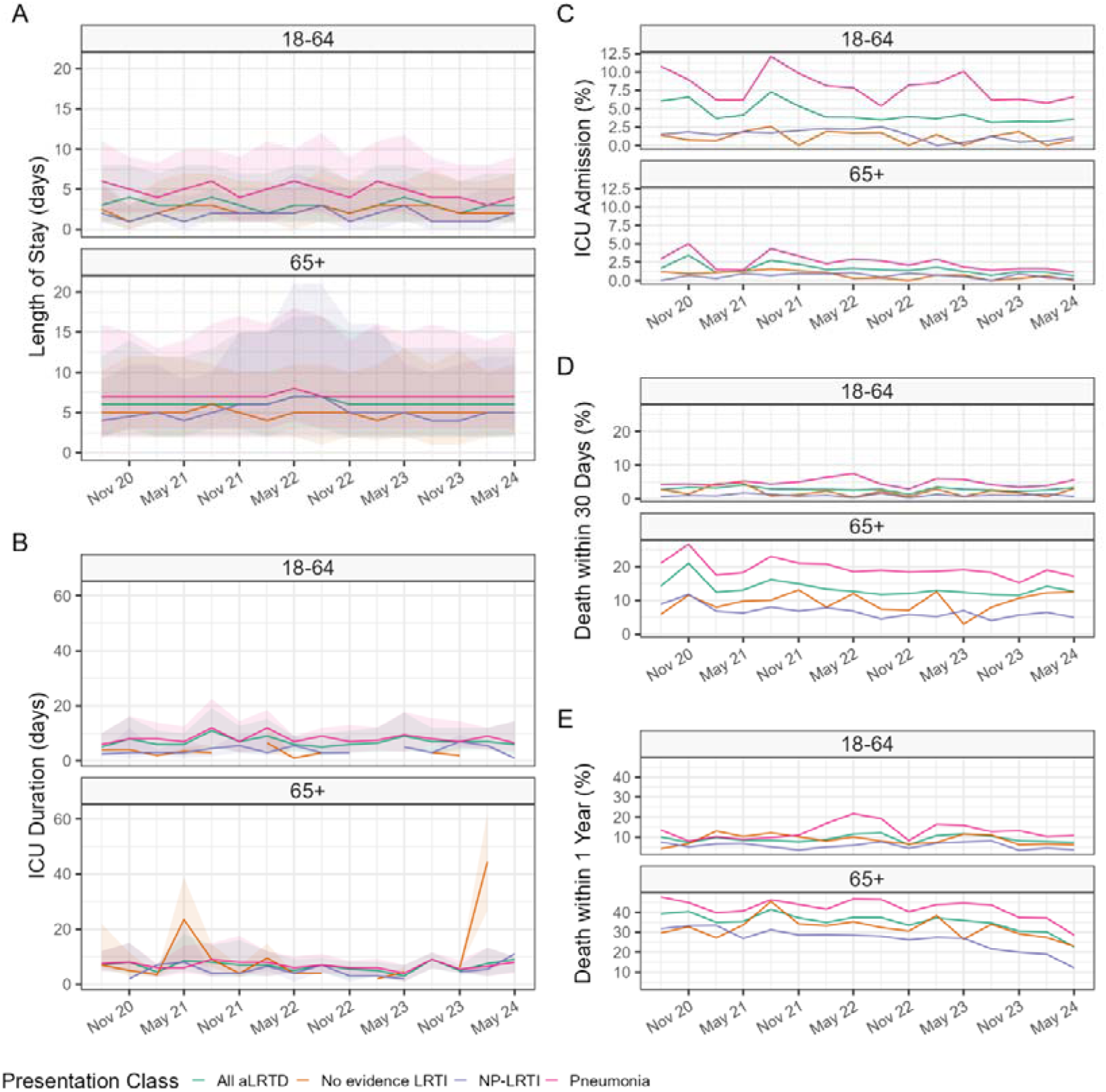
Quarterly Outcomes of aLTRD hospitalisations in different clinical presentations between Aug 2020 and Jul 2024 by Age Group. (A)Quarterly length of hospital stay in days, median(lines) and interquartile range(shaded areas) stratified by age group (B)Quarterly length of ICU admission in days, median(lines) and interquartile range(shaded areas) stratified by age group (C) Quarterly percentage of confirmed ICU admissions stratified by age group (D) Quarterly percentage of confirmed deaths within 30 days stratified by age group (E) Quarterly percentage of confirmed deaths within 1 year stratified by age group; stratified by clinical presentation for pneumonia, non-pneumonic lower respiratory tract infection (NP-LRTI) and cases with no evidence of lower respiratory tract infection (LRTI) *Only individuals with complete follow-up (1 year for mortality within 1 year; 30 days for mortality within 30 days) were included.

In the Cox proportional hazard model for death within 30 days of admission, relative to patients with pneumonia, patients with NP-LRTI had a significantly lower hazard ratio (HR) of 0.32 (95% CI: 0.29 – 0.35) and those with no evidence of LRTI had an HR of 0.43 (95% CI: 0.19 – 0.97). Age was a significant factor, with increasing age associated with higher hazard ratios: patients aged 85+ had an HR of 5.1 (95% CI: 2.8 – 9.5) compared to the 18-34 age group, and men had a higher risk of death compared to women (HR: 1.2, 95% CI: 1.1-1.3). Patients with heart failure also had a higher risk (HR: 1.3, 95% CI: 1.2 – 1.4) while the presence of chronic respiratory disease exacerbation was associated with a lower risk of death (HR: 0.9, 95% CI: 0.84 – 0.95). Severe comorbidities, as indicated by the CCI, were significantly associated with mortality, having an HR of 3.4 (95% CI: 2.8 – 4.1). Over successive study years, the hazard ratios for death within 30 days decreased, suggesting improvements in patient outcomes over time (Table 3). For hospital discharge within 30 days, based on the adjusted survival analysis, patients with NP-LRTI had an increased likelihood of discharge than those with pneumonia (HR: 1.5, 95% CI: 1.5 – 1.5), as did those with no evidence of LRTI (HR: 1.4, 95% CI: 1.1 – 1.8). Younger age groups were more likely to be discharged within 30 days, with the likelihood decreasing as age increased. Patients with heart failure, vs those without, were less likely to be discharged within 30 days (HR: 0.69, 95% CI: 0.67 – 0.71). Severe comorbidities (CCI >4), vs those without, were also associated with a lower likelihood of discharge (HR: 0.73, 95% CI: 0.7 – 0.77). Over the study period, discharge likelihood gradually increased (Table 3). Kaplan-Meier curves illustrating these outcomes are in Supplementary Data S11.

**Table 3:**
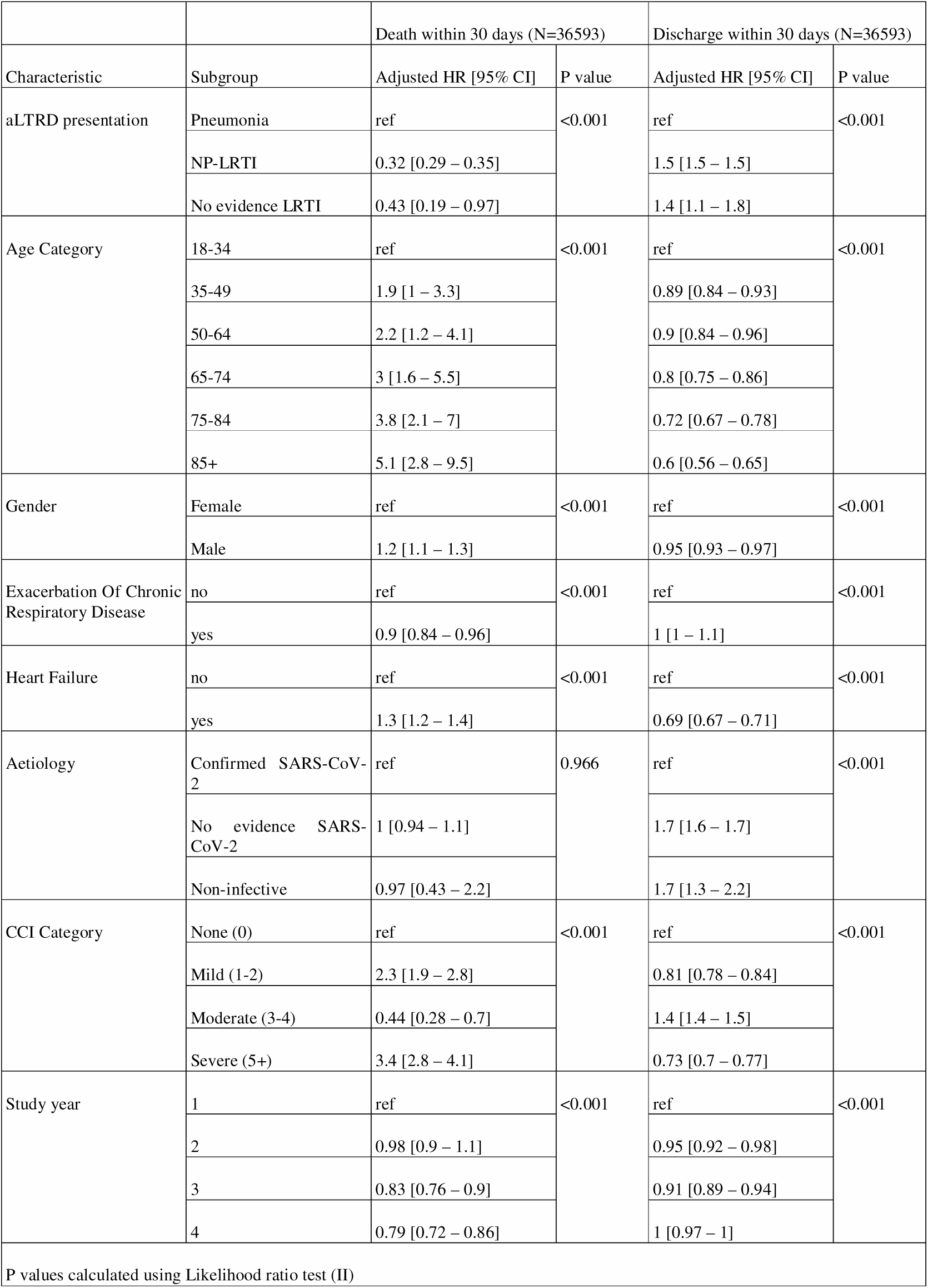
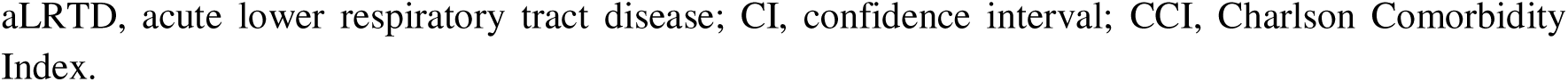
Adjusted Cox proportional hazards models for death and hospital discharge following admission with aLRTD. The outcome of death within 30 days is censored at the earliest of 30 days or 31^st^ July 2024; the outcome of discharge is censored at 30 days or death. In the analysis for death, an increased hazard implies a poorer outcome; whereas, in the analysis for hospital discharge, an increased probability of discharge is equivalent to a reduced length of stay, and lower hazard rate implies a poorer outcome.

## DISCUSSION

This four-year, two-site single-centre study conducted within a defined geographical area provides a comprehensive overview of aLRTD during the COVID-19 pandemic and the subsequent post-pandemic period. It is the first UK study to report such an analysis, thoroughly monitoring all aLRTD cases to determine the incidence and severity of different clinical presentations in hospitalised individuals: pneumonia; NP-LRTI and aLRTD with no evidence of LRTI.

Our findings indicate that the burden of aLRTD remains substantial. While COVID-19 significantly impacted aLRTD incidence rates, particularly in the initial years of the pandemic, non-COVID infections remained consistent The estimates compare to a retrospective analysis of 2018-19 hospitalisations, showing an annual incidence of aLRTD of 1901 per 100,000 adults, pneumonia 591/100,000 and NP-LRTI 739/100,000^13^ and also aligns with pre-pandemic estimates from other countries^14,15^. However, the 638/100,000 pneumonia incidence observed here (2020-24) was higher than Nottingham’s 96.3/100,000 (2013-18)^16^. Methodological differences likely explain this, including comprehensive medical record-based surveillance and a study-specific population catchment analysis ensuring a precise incidence denominator^8^. Differences in case definitions and potential under-ascertainment of pneumonia events in Nottingham are supported by other UK data^17^, which indicated lower pneumococcal CAP incidence estimates compared to national respiratory illness admission data. The stability in non-COVID aLRTD incidence we found suggests that the underlying burden of respiratory diseases persisted despite fluctuations caused by the pandemic. Pneumonia was the most common aLRTD presentation, with higher incidence rates than NP-LRTI, as we previously reported in 2022 and 2020^5,13^. However, NP-LRTI still represented a considerable proportion of cases, highlighting the importance of including NP-LRTI in epidemiological studies and public health strategies to avoid underestimating the overall disease burden.

Throughout the study, pneumonia and NP-LRTI admissions varied significantly, driven by confirmed SARS-CoV-2 cases, mirroring the incidence of COVID-19 in the community^18^ affected by periods of non-pharmaceutical interventions in the UK^19^, the emergence of variants^18^, seasonal respiratory infection patterns, and the timing of vaccination campaigns. The Alpha variant (B.1.1.7) contributed to increased pneumonia incidence due to its severity^20,21^, while Delta’s (B.1.617.2) potential impact was moderated by ongoing restrictions and the rollout of vaccination^22^. The proportion of pneumonia cases can be considered as an indirect measure of disease severity in respiratory infections. Omicron (B.1.1.529) caused a surge in infections, but the associated proportion of pneumonia cases declined, consistent with its reduced severity compared to previous variants, and the population’s enhanced immunity through vaccination. This trend was especially notable in older age groups, where the combination of vaccination and prior exposure contributed to reduced disease severity, even during periods of high transmission. The persistently lower incidence rates of NP-LRTI during the latter half of 2020 and early 2021 highlight the potential residual impact of public health measures on respiratory infections beyond COVID-19. However, in contrast to previous studies^23^, we did not find a decline in hospital admissions for either pneumonia or NP-LRTI unrelated to COVID-19 when NPI measures were implemented to control the pandemic. Finally, the post-2022 pattern may indicate a shift toward endemic COVID-19 and the re-establishment of regular seasonal cycles for other respiratory diseases.

Clinical outcomes of aLRTD varied by clinical presentation. Pneumonia patients had worse outcomes, including longer hospital stays, higher ICU admission rates, prolonged ICU durations, and increased mortality, highlighting the severity of pneumonia. Older adults experienced longer hospital stays reflecting their increased burden and slower recovery rates. Length of stay remained stable during the pandemic and in the subsequent years, suggesting consistent treatment approaches. ICU admissions spiked during the Delta wave due to its higher transmissibility and severity^24^, then decreased with the less severe Omicron variant^22^. Younger patients were prioritized for ICU care^25^, while older patients were less likely to be admitted due to limited benefits and potential risks^26^.

Cox model analysis provided additional insights into the factors influencing mortality and discharge outcomes for aLRTD patients: there was a significant reduction in the risk of death associated with non-pneumonic LRTD and men were found to have a higher risk of death compared to women. Interestingly, while CRDE did not increase the risk of death, heart failure did confer additional risk. Although potentially surprising, the former finding aligns with previous research^27^ on COPD outcomes when severity was considered. Over the four-year period, the severity of COVID-19, reflected in the risk of death within 30 days, was comparable to other infectious causes, most likely due to the widespread vaccination uptake. Regarding discharge outcomes, patients with pneumonic aLRTD had a longer length of stay, indicating more severe disease. Additionally, COVID-19 remained associated with a longer length of hospital stay, aligning with findings from studies on the Delta and Omicron variants^22^.

This study had several strengths. Firstly, it was conducted prospectively with active screening of hospital admissions for aLRTD symptoms, allowing for comprehensive, case-by-case assessment in a well-defined geographic area. Unlike studies relying on ICD-10 coding or national data-linkage, this approach enabled detailed, individualised data collection. Secondly, by using consultee consent and specific authorization ensured full representation of all hospitalized adults, including those lacking capacity. Conducted across two hospitals providing all acute secondary care for Bristol, this design ensured comprehensive surveillance in a defined population, yielding accurate estimates of disease incidence and severity. Additionally, linking medical and community records allowed for detailed data capture; incidence calculations based on GP records and hospital utilization data provided greater accuracy than estimates based on geographic population boundaries and census data.

There were several limitations to this study. First, while the study was conducted over four years and across two hospitals in Bristol, the findings may not be fully generalisable to other regions, or healthcare systems. The cohort was predominantly of White-British ethnicity, limiting applicability to more diverse populations. Changes in healthcare access and patient behavior during the pandemic could have impacted admission rates and severity assessments. Although the study hospitals maintained acute care capacity, these potential behavioural shifts represent an important consideration.

In conclusion, our study revealed the enduring burden of aLRTD during and after the COVID-19 pandemic. While hospital admissions for pneumonia and NP-LRTI initially showed significant variability due to SARS-CoV-2, post-2022 patterns indicated a shift towards endemic COVID-19 and a return to seasonal respiratory illness cycles. The data we present re-emphasises the importance of advancing strategies to prevent aLRTD through immunisation. Recent improvements in vaccines against influenza, COVID-19, RSV and pneumococcal infections need to be supplemented by improved antimicrobial treatments and further advances in immunisation against these and other respiratory pathogens.

## Data Availability

The data used in this study are sensitive and cannot be made publicly available without breaching patient confidentiality rules. Therefore, individual participant data and a data dictionary are not available to other researchers.

## ACKNOWLEDGEMENTS

We thank colleagues at the University of Bristol for their support with this study, including Rachel Davies, Paul Savage, Emma Foose, Susan Christie, Mark Mummé, Alison Horne, Mai Baquedano, and Adam Taylor. We would like to thank Stewart Robinson, David Clint and Henry Stuart and their teams for the support provided during this study. We would also like to acknowledge the research teams at North Bristol and University Hospitals of Bristol and Weston NHS Trusts for making this study possible, including Helen Lewis-White, Rebecca Smith, Rajeka Lazarus, Jane Blazeby, Diana Benton, and David Wynick. Finally, we would like to thank Aman Kaur-Singh and Kevin Sweetland for their support with this study.

## AUTHORSHIP STATEMENT

CH, AC, RC, ML, EB, JS, BG, AF, LD generated the research questions and analysis plans. The AvonCAP research team, MC, SM and CH were involved in data collection. CH, AC, RC, LD and AF undertook data analysis. CH, AC, RC, ML, EB, SM, MC, JS, JC, JO, GE, NM, BG, LD, AF contributed to preparation of the manuscript and its revisions before publication. CH and AF provided oversight of the research.

## DATA SHARING

### The AvonCAP Research Group

Aaran Sinclair, Alessandra Mantini, Alexander Tremaine, Alison Horne, Amelia Langdon, Amy Taylor, Anabella Turner, Anna Jones, Anna Morley, Anya Mattocks, Archana Sudharsanan, Bethany Osborne, Brianna Dooley, Claire Mitchell, Emma Bridgeman, Emma Dickason-Palmer, Emma Scott, Ffion Davies, Fiona Perkins, Francesca Bayley, Francis Mensah, Gabriella Ruffino, Gabriella Valentine, Genevive Coulter, Grace Tilzey, Harriet Ibbotson, Jane Kinney, Johanna Kellett Wright, Joseph Cleere, Josephine Bonnici, Juan Garcia Tello Julia Brzezinska Julie Cloake, Katarina Milutinovic, Kate Helliker, Katie Maughan, Kazminder Fox, Konstantina Minou, Lana Ward, Laura Jaramillo, Leah Fleming, Leigh Morrison, Lily Smart, Lisa Grimmer, Louise Wright, Lucy Grimwood, Maddalena Bellavia, Maddie Clout, Mai Baquedano, Maise Borril, Maria Garcia Gonzalez, Marianne Vasquez, Martina Chmelarova, Matthew Jepson, Milo Jeenes-Flanagan, Natalie Chang, Nefeli Taravira, Niall Grace, Nicola Manning, Oliver Griffiths, Peter Sequenza, Pip Croxford, Rajeka Lazarus, Rebecca Abou Chullieh, Rebecca Clemence, Rhian Walters, Riley Cooper, Robert Graham, Robin Marlow, Robyn Heath, Rupert Antico, Sandi Nammuni Arachchge, Sarah Jenkins, Sean Robinson, Seevakumar Suppiah, Serena McGuinness, Siddiqa Uddin, Taslima Mona, Tawassal Riaz, Thomas Wright, Vicki Mackay, Zandile Maseko, Zoe Taylor, Zsolt Friedrich, Zsuzsa Szasz-Benczur

