## Supplementary Data for "Incidence of community acquired lower respiratory tract disease in Bristol, UK following the emergence of SARS-CoV-2: a prospective cohort study 2020-2024"

**Supplementary Data 1 (S1): Characteristics of patients hospitalised with all aLRTD for the period Aug 2020 to Jul 24 by study year**

|  |  | All aLRTD | Year 1, 2020-21 | Year 2, 2021-22 | Year 3, 2022-23 | Year 4, 2023-24 |  |
| --- | --- | --- | --- | --- | --- | --- | --- |
| Variable | Characteristic | Value (N=42834) | Value (N=9583) | Value (N=11777) | Value (N=10890) | Value (N=10584) | P value |
| Age | Median [IQR] | 73.3 [57.7—83.3] | 73.1 [57.4—83.3] | 71.9 [54.7—82.6] | 74.3 [60.1—83.9] | 73.8 [58.2—83.4] | <0.001 † |
| Age Category | 18-34 % (n) | 7.5% (3210) | 6.3% (600) | 9.7% (1145) | 6.4% (697) | 7.3% (768) | <0.001 †† |
|  | 35-49 % (n) | 9.9% (4244) | 10.5% (1010) | 11.0% (1300) | 8.6% (932) | 9.5% (1002) |  |
|  | 50-64 % (n) | 17.9% (7667) | 18.8% (1803) | 17.9% (2110) | 17.0% (1847) | 18.0% (1907) |  |
|  | 65-74 % (n) | 18.9% (8110) | 19.3% (1850) | 18.4% (2168) | 19.8% (2158) | 18.3% (1934) |  |
|  | 75-84 % (n) | 24.9% (10678) | 24.1% (2313) | 23.3% (2743) | 26.2% (2848) | 26.2% (2774) |  |
|  | 85+ % (n) | 20.8% (8925) | 20.9% (2007) | 19.6% (2311) | 22.1% (2408) | 20.8% (2199) |  |
| Age Eligible for Pneumococcal vaccination | 18-64 % (n) | 35.3% (15121) | 35.6% (3413) | 38.7% (4555) | 31.9% (3476) | 34.7% (3677) | <0.001 †† |
|  | 65+ % (n) | 64.7% (27713) | 64.4% (6170) | 61.3% (7222) | 68.1% (7414) | 65.3% (6907) |  |
| Gender | Female % (n) | 50.6% (21683) | 49.4% (4736) | 50.0% (5890) | 51.3% (5588) | 51.7% (5469) | 0.0027 †† |
| Ethnicity | White British % (n) | 75.3% (32264) | 78.1% (7484) | 71.4% (8411) | 74.6% (8128) | 77.9% (8241) | <0.001 †† |
|  | White other % (n) | 2.9% (1223) | 2.6% (252) | 3.2% (378) | 2.5% (275) | 3.0% (318) |  |
|  | Mixed origin % (n) | 1.0% (435) | 0.9% (82) | 1.0% (118) | 1.2% (127) | 1.0% (108) |  |
|  | Black % (n) | 1.9% (826) | 1.8% (168) | 2.1% (250) | 1.9% (207) | 1.9% (201) |  |
|  | Asian % (n) | 2.1% (912) | 2.9% (280) | 2.0% (237) | 1.7% (183) | 2.0% (212) |  |
|  | Other race % (n) | 1.1% (487) | 0.9% (88) | 1.2% (147) | 0.9% (97) | 1.5% (155) |  |
|  | Unknown % (n) | 15.6% (6686) | 12.8% (1229) | 19.0% (2235) | 17.2% (1873) | 12.7% (1349) |  |
|  | <missing> % (n) | 0.0% (1) | 0.0% (0) | 0.0% (1) | 0.0% (0) | 0.0% (0) |  |
| Care Home Resident | yes % (n) | 8.2% (3513) | 10.1% (966) | 6.7% (789) | 8.2% (892) | 8.2% (866) | <0.001 †† |
| Smoker | Non-smoker % (n) | 36.7% (15733) | 36.0% (3447) | 39.1% (4606) | 35.0% (3810) | 36.6% (3870) | <0.001 †† |
|  | Current % (n) | 12.1% (5168) | 10.0% (955) | 10.0% (1178) | 14.1% (1540) | 14.1% (1495) |  |
|  | Ex-smoker % (n) | 43.1% (18470) | 42.2% (4042) | 43.6% (5136) | 44.4% (4839) | 42.1% (4453) |  |
|  | Unknown % (n) | 8.1% (3463) | 11.9% (1139) | 7.3% (857) | 6.4% (701) | 7.2% (766) |  |
| Heart Failure | yes % (n) | 18.6% (7952) | 20.6% (1975) | 16.7% (1971) | 18.5% (2018) | 18.8% (1988) | <0.001 †† |
| Exacerbation Of Chronic Respiratory Disease | yes % (n) | 44.9% (19252) | 42.8% (4102) | 40.6% (4780) | 47.7% (5196) | 48.9% (5174) | <0.001 †† |
| CCI Category | None (0) % (n) | 14.3% (6145) | 16.0% (1535) | 15.3% (1799) | 12.6% (1368) | 13.6% (1443) | <0.001 †† |
|  | Mild (1-2) % (n) | 27.3% (11679) | 26.6% (2552) | 27.0% (3178) | 28.9% (3147) | 26.5% (2802) |  |
|  | Moderate (3-4) % (n) | 13.2% (5654) | 12.4% (1189) | 16.3% (1923) | 11.1% (1214) | 12.5% (1328) |  |
|  | Severe (5+) % (n) | 45.2% (19356) | 44.9% (4307) | 41.4% (4877) | 47.4% (5161) | 47.3% (5011) |  |
| CURB 65 Category | 0-1 (Mild) % (n) | 74.9% (32102) | 72.6% (6958) | 75.2% (8858) | 73.9% (8052) | 77.8% (8234) | <0.001 †† |
|  | 2 (Moderate) % (n) | 21.6% (9267) | 22.3% (2138) | 21.0% (2471) | 23.4% (2543) | 20.0% (2115) |  |
|  | 3-5 (Severe) % (n) | 3.4% (1465) | 5.1% (487) | 3.8% (448) | 2.7% (295) | 2.2% (235) |  |
| COPD | yes % (n) | 28.0% (12006) | 27.0% (2591) | 25.0% (2941) | 30.8% (3356) | 29.5% (3118) | <0.001 †† |
| Asthma | yes % (n) | 16.6% (7090) | 15.9% (1523) | 15.4% (1814) | 16.3% (1774) | 18.7% (1979) | <0.001 †† |
| Bronchiectasis | no % (n) | 95.6% (40968) | 96.2% (9221) | 96.2% (11333) | 95.4% (10387) | 94.7% (10027) | <0.001 †† |
|  | yes % (n) | 4.3% (1863) | 3.8% (362) | 3.7% (441) | 4.6% (503) | 5.3% (557) |  |
|  | <missing> % (n) | 0.0% (3) | 0.0% (0) | 0.0% (3) | 0.0% (0) | 0.0% (0) |  |
| IHD | no % (n) | 86.3% (36969) | 85.6% (8203) | 87.6% (10320) | 86.6% (9426) | 85.2% (9020) | <0.001 †† |
|  | yes % (n) | 13.7% (5864) | 14.4% (1380) | 12.4% (1457) | 13.4% (1464) | 14.8% (1563) |  |
|  | <missing> % (n) | 0.0% (1) | 0.0% (0) | 0.0% (0) | 0.0% (0) | 0.0% (1) |  |
| Any Immune Compromise | yes % (n) | 13.1% (5618) | 8.6% (821) | 10.1% (1194) | 15.7% (1712) | 17.9% (1891) | <0.001 †† |
| Diabetes Type | None % (n) | 79.6% (34075) | 77.9% (7466) | 80.8% (9510) | 79.2% (8630) | 80.0% (8469) | <0.001 †† |
|  | Type 1 % (n) | 1.2% (522) | 1.2% (113) | 1.3% (150) | 1.3% (145) | 1.1% (114) |  |
|  | Type 2 % (n) | 19.2% (8236) | 20.9% (2004) | 18.0% (2117) | 19.4% (2115) | 18.9% (2000) |  |
|  | <missing> % (n) | 0.0% (1) | 0.0% (0) | 0.0% (0) | 0.0% (0) | 0.0% (1) |  |
| CKD | None % (n) | 75.1% (32149) | 76.1% (7296) | 77.5% (9123) | 72.4% (7882) | 74.1% (7848) | <0.001 †† |
|  | Mild (CKD 1-3) % (n) | 21.2% (9064) | 19.8% (1898) | 19.2% (2258) | 23.5% (2559) | 22.2% (2349) |  |
|  | Moderate or Severe CKD (CKD 4+) % (n) | 3.8% (1620) | 4.1% (389) | 3.4% (396) | 4.1% (449) | 3.6% (386) |  |
|  | <missing> % (n) | 0.0% (1) | 0.0% (0) | 0.0% (0) | 0.0% (0) | 0.0% (1) |  |
| Pneumococcal vaccination | Not received % (n) | 43.5% (18642) | 39.9% (3821) | 40.9% (4821) | 46.5% (5063) | 46.6% (4937) | <0.001 †† |
|  | PPV23 % (n) | 47.9% (20535) | 51.5% (4940) | 49.2% (5800) | 45.7% (4977) | 45.5% (4818) |  |
|  | PCV13 % (n) | 0.4% (180) | 0.0% (0) | 0.2% (26) | 1.1% (121) | 0.3% (33) |  |
|  | Unknown % (n) | 8.1% (3477) | 8.6% (822) | 9.6% (1130) | 6.7% (729) | 7.5% (796) |  |
| Influenza Vaccination | Not received % (n) | 31.5% (13473) | 35.0% (3354) | 29.6% (3486) | 28.3% (3087) | 33.5% (3546) | <0.001 †† |
|  | Received % (n) | 63.2% (27066) | 56.6% (5422) | 61.2% (7211) | 69.5% (7571) | 64.8% (6862) |  |
|  | Unknown % (n) | 5.3% (2282) | 8.4% (807) | 9.2% (1080) | 2.1% (232) | 1.5% (163) |  |
|  | <missing> % (n) | 0.0% (13) | 0.0% (0) | 0.0% (0) | 0.0% (0) | 0.1% (13) |  |
| †, Kruskal-Wallis rank sum test (continuous); ††, Fisher's exact test (categorical)  Normal distributions determined by the Anderson-Darling test (P>0.005)  An adjusted P value of 0.0025 may be considered significant. | | | | | | |  |

In the UK, patients aged ≥65 years are eligible for Pneumococcal vaccination (PneumoVax®, PPV23) once, and annual influenza vaccine (please see Green Book23.

*Chronic kidney disease (CKD) was classified as mild if stage 1-3; moderate/severe if stage 4-5, end-stage renal failure or there was dialysis dependence.

**Influenza vaccination indicates vaccinated in the last 12 months.

aLRTD, acute lower respiratory tract disease; CCI, Charlson Comorbidity Index; CKD, chronic kidney disease; COPD, chronic obstructive pulmonary disease; IHD, ischaemic heart disease; IQR, interquartile range; PCV13, pneumococcal conjugate vaccine (13-valent); PPV23 pneumococcal polysaccharide vaccine (23-valent)

**Supplementary Data 2 (S2): Characteristics of patients hospitalised with Pneumonia for the period Aug 2020 to Jul 24 by study year**

|  |  | Pneumonia | Year 1, 2020-21 | Year 2, 2021-22 | Year 3, 2022-23 | Year 4, 2023-24 |  |
| --- | --- | --- | --- | --- | --- | --- | --- |
| Variable | Characteristic | Value (N=20639) | Value (N=5285) | Value (N=5108) | Value (N=4979) | Value (N=5267) | P value |
| Age | Median [IQR] | 74.5 [59.5—84.1] | 72.7 [56.9—83.1] | 73.7 [58.3—83.8] | 75.9 [63.5—84.9] | 75.4 [60.3—84.6] | <0.001 † |
| Age Category | 18-34 % (n) | 5.1% (1055) | 5.1% (267) | 5.7% (293) | 4.0% (201) | 5.6% (294) | <0.001 †† |
|  | 35-49 % (n) | 9.6% (1976) | 11.2% (591) | 10.7% (548) | 7.4% (370) | 8.9% (467) |  |
|  | 50-64 % (n) | 18.1% (3738) | 20.9% (1106) | 18.3% (935) | 15.8% (786) | 17.3% (911) |  |
|  | 65-74 % (n) | 18.6% (3829) | 18.3% (966) | 19.0% (972) | 19.7% (979) | 17.3% (912) |  |
|  | 75-84 % (n) | 26.0% (5356) | 23.9% (1262) | 24.5% (1249) | 28.3% (1411) | 27.2% (1434) |  |
|  | 85+ % (n) | 22.7% (4685) | 20.7% (1093) | 21.8% (1111) | 24.7% (1232) | 23.7% (1249) |  |
| Age Eligible for Pneumococcal vaccination | 18-64 % (n) | 32.8% (6769) | 37.2% (1964) | 34.8% (1776) | 27.3% (1357) | 31.7% (1672) | <0.001 †† |
|  | 65+ % (n) | 67.2% (13870) | 62.8% (3321) | 65.2% (3332) | 72.7% (3622) | 68.3% (3595) |  |
| Gender | Female % (n) | 47.4% (9780) | 45.5% (2404) | 45.6% (2330) | 49.5% (2465) | 49.0% (2581) | <0.001 †† |
| Ethnicity | White British % (n) | 75.5% (15575) | 76.8% (4060) | 70.3% (3593) | 76.2% (3793) | 78.4% (4129) | <0.001 †† |
|  | White other % (n) | 3.0% (621) | 2.9% (151) | 3.7% (189) | 2.3% (115) | 3.2% (166) |  |
|  | Mixed origin % (n) | 0.8% (173) | 0.8% (44) | 0.7% (34) | 0.8% (40) | 1.0% (55) |  |
|  | Black % (n) | 2.0% (406) | 2.2% (114) | 2.2% (110) | 1.8% (91) | 1.7% (91) |  |
|  | Asian % (n) | 2.2% (448) | 3.7% (195) | 2.0% (101) | 1.4% (68) | 1.6% (84) |  |
|  | Other race % (n) | 1.1% (234) | 1.0% (54) | 1.2% (60) | 0.8% (42) | 1.5% (78) |  |
|  | Unknown % (n) | 15.4% (3182) | 12.6% (667) | 20.0% (1021) | 16.7% (830) | 12.6% (664) |  |
| Care Home Resident | yes % (n) | 10.0% (2054) | 11.3% (598) | 8.3% (422) | 9.9% (491) | 10.3% (543) | <0.001 †† |
| Smoker | Non-smoker % (n) | 37.0% (7639) | 37.5% (1980) | 39.5% (2018) | 34.6% (1721) | 36.5% (1920) | <0.001 †† |
|  | Current % (n) | 11.0% (2264) | 8.7% (462) | 8.8% (451) | 13.6% (679) | 12.8% (672) |  |
|  | Ex-smoker % (n) | 43.8% (9045) | 41.6% (2197) | 45.0% (2297) | 44.9% (2238) | 43.9% (2313) |  |
|  | Unknown % (n) | 8.2% (1691) | 12.2% (646) | 6.7% (342) | 6.8% (341) | 6.9% (362) |  |
| Heart Failure | yes % (n) | 16.8% (3474) | 16.2% (854) | 14.3% (728) | 19.2% (958) | 17.7% (934) | <0.001 †† |
| Exacerbation Of Chronic Respiratory Disease | yes % (n) | 42.2% (8711) | 39.2% (2070) | 38.3% (1954) | 46.4% (2310) | 45.1% (2377) | <0.001 †† |
| CCI Category | None (0) % (n) | 14.7% (3031) | 18.1% (959) | 15.5% (793) | 12.1% (602) | 12.9% (677) | <0.001 †† |
|  | Mild (1-2) % (n) | 27.2% (5622) | 26.9% (1423) | 27.7% (1413) | 28.2% (1406) | 26.2% (1380) |  |
|  | Moderate (3-4) % (n) | 10.7% (2207) | 11.8% (623) | 12.3% (626) | 7.8% (389) | 10.8% (569) |  |
|  | Severe (5+) % (n) | 47.4% (9779) | 43.1% (2280) | 44.6% (2276) | 51.9% (2582) | 50.1% (2641) |  |
| CURB 65 Category | 0-1 (Mild) % (n) | 71.4% (14738) | 71.7% (3788) | 70.3% (3591) | 69.5% (3459) | 74.0% (3900) | <0.001 †† |
|  | 2 (Moderate) % (n) | 24.3% (5009) | 22.9% (1210) | 24.4% (1245) | 27.0% (1346) | 22.9% (1208) |  |
|  | 3-5 (Severe) % (n) | 4.3% (892) | 5.4% (287) | 5.3% (272) | 3.5% (174) | 3.0% (159) |  |
| COPD | yes % (n) | 26.8% (5538) | 24.2% (1280) | 24.0% (1226) | 31.2% (1554) | 28.1% (1478) | <0.001 †† |
| Asthma | yes % (n) | 13.9% (2877) | 14.2% (750) | 12.9% (659) | 13.3% (664) | 15.3% (804) | 0.0029 †† |
| Bronchiectasis | no % (n) | 95.1% (19628) | 96.3% (5087) | 95.9% (4899) | 94.3% (4696) | 93.9% (4946) | <0.001 †† |
|  | yes % (n) | 4.9% (1009) | 3.7% (198) | 4.1% (207) | 5.7% (283) | 6.1% (321) |  |
|  | <missing> % (n) | 0.0% (2) | 0.0% (0) | 0.0% (2) | 0.0% (0) | 0.0% (0) |  |
| IHD | yes % (n) | 13.5% (2796) | 13.3% (704) | 13.1% (669) | 12.6% (629) | 15.1% (794) | 0.0019 †† |
| Any Immune Compromise | yes % (n) | 12.4% (2564) | 7.8% (411) | 10.3% (528) | 15.4% (768) | 16.3% (857) | <0.001 †† |
| Diabetes Type | None % (n) | 79.3% (16360) | 78.3% (4136) | 79.6% (4068) | 78.9% (3930) | 80.2% (4226) | 0.035 †† |
|  | Type 1 % (n) | 1.1% (227) | 1.0% (54) | 1.2% (63) | 1.3% (66) | 0.8% (44) |  |
|  | Type 2 % (n) | 19.6% (4052) | 20.7% (1095) | 19.1% (977) | 19.7% (983) | 18.9% (997) |  |
| CKD | None % (n) | 74.1% (15287) | 76.8% (4057) | 75.7% (3866) | 70.6% (3516) | 73.1% (3848) | <0.001 †† |
|  | Mild (CKD 1-3) % (n) | 21.9% (4526) | 19.4% (1024) | 20.6% (1052) | 25.1% (1248) | 22.8% (1202) |  |
|  | Moderate or Severe CKD (CKD 4+) % (n) | 4.0% (826) | 3.9% (204) | 3.7% (190) | 4.3% (215) | 4.1% (217) |  |
| Pneumococcal vaccination | Not received % (n) | 42.2% (8704) | 41.7% (2204) | 38.6% (1973) | 43.8% (2182) | 44.5% (2345) | <0.001 †† |
|  | PPV23 % (n) | 49.0% (10111) | 48.5% (2562) | 51.4% (2626) | 48.0% (2392) | 48.1% (2531) |  |
|  | PCV13 % (n) | 0.4% (77) | 0.0% (0) | 0.3% (13) | 1.0% (52) | 0.2% (12) |  |
|  | Unknown % (n) | 8.5% (1747) | 9.8% (519) | 9.7% (496) | 7.1% (353) | 7.2% (379) |  |
| Influenza Vaccination | Not received % (n) | 31.2% (6435) | 35.9% (1895) | 29.8% (1522) | 26.9% (1338) | 31.9% (1680) | <0.001 †† |
|  | Received % (n) | 63.3% (13062) | 54.5% (2879) | 61.0% (3115) | 71.5% (3560) | 66.6% (3508) |  |
|  | Unknown % (n) | 5.5% (1137) | 9.7% (511) | 9.2% (471) | 1.6% (81) | 1.4% (74) |  |
|  | <missing> % (n) | 0.0% (5) | 0.0% (0) | 0.0% (0) | 0.0% (0) | 0.1% (5) |  |
| †, Kruskal-Wallis rank sum test (continuous); ††, Fisher's exact test (categorical)  Normal distributions determined by the Anderson-Darling test (P>0.005)  An adjusted P value of 0.0025 may be considered significant. | | | | | | |  |

In the UK, patients aged ≥65 years are eligible for Pneumococcal vaccination (PneumoVax®, PPV23) once, and annual influenza vaccine (please see Green Book23.

*Chronic kidney disease (CKD) was classified as mild if stage 1-3; moderate/severe if stage 4-5, end-stage renal failure or there was dialysis dependence.

**Influenza vaccination indicates vaccinated in the last 12 months.

aLRTD, acute lower respiratory tract disease; CCI, Charlson Comorbidity Index; CKD, chronic kidney disease; COPD, chronic obstructive pulmonary disease; IHD, ischaemic heart disease; IQR, interquartile range; PCV13, pneumococcal conjugate vaccine (13-valent); PPV23 pneumococcal polysaccharide vaccine (23-valent)

**Supplementary Data 3 (S3): Characteristics of patients hospitalised with NP-LRTI for the period Aug 2020 to Jul 24 by study year**

|  |  | NP-LRTI | Year 1, 2020-21 | Year 2, 2021-22 | Year 3, 2022-23 | Year 4, 2023-24 |  |
| --- | --- | --- | --- | --- | --- | --- | --- |
| Variable | Characteristic | Value (N=15061) | Value (N=2547) | Value (N=4760) | Value (N=4234) | Value (N=3520) | P value |
| Age | Median [IQR] | 71 [53—81.7] | 71.5 [53.7—81.6] | 68.8 [47.9—80.8] | 72.6 [56.5—82.8] | 70.9 [54.4—81.8] | <0.001 † |
| Age Category | 18-34 % (n) | 11.3% (1695) | 9.7% (248) | 14.7% (698) | 9.4% (396) | 10.0% (353) | <0.001 †† |
|  | 35-49 % (n) | 11.1% (1669) | 12.2% (312) | 12.0% (572) | 9.7% (410) | 10.7% (375) |  |
|  | 50-64 % (n) | 17.7% (2672) | 16.5% (420) | 17.8% (845) | 17.5% (742) | 18.9% (665) |  |
|  | 65-74 % (n) | 18.7% (2823) | 19.9% (506) | 17.2% (820) | 19.7% (833) | 18.9% (664) |  |
|  | 75-84 % (n) | 23.2% (3492) | 23.8% (605) | 21.3% (1015) | 23.6% (998) | 24.8% (874) |  |
|  | 85+ % (n) | 18.0% (2710) | 17.9% (456) | 17.0% (810) | 20.2% (855) | 16.7% (589) |  |
| Age Eligible for Pneumococcal vaccination | 18-64 % (n) | 40.1% (6036) | 38.5% (980) | 44.4% (2115) | 36.6% (1548) | 39.6% (1393) | <0.001 †† |
|  | 65+ % (n) | 59.9% (9025) | 61.5% (1567) | 55.6% (2645) | 63.4% (2686) | 60.4% (2127) |  |
| Gender | Female % (n) | 54.1% (8149) | 54.5% (1389) | 53.9% (2567) | 53.3% (2256) | 55.0% (1937) | 0.46 †† |
| Ethnicity | White British % (n) | 74.5% (11223) | 81.1% (2066) | 71.3% (3393) | 72.1% (3052) | 77.0% (2712) | <0.001 †† |
|  | White other % (n) | 2.7% (413) | 2.4% (60) | 2.9% (137) | 2.8% (119) | 2.8% (97) |  |
|  | Mixed origin % (n) | 1.2% (188) | 1.0% (26) | 1.3% (61) | 1.4% (61) | 1.1% (40) |  |
|  | Black % (n) | 1.9% (284) | 1.4% (36) | 2.2% (104) | 1.7% (74) | 2.0% (70) |  |
|  | Asian % (n) | 2.3% (341) | 2.4% (60) | 2.0% (97) | 2.1% (88) | 2.7% (96) |  |
|  | Other race % (n) | 1.2% (184) | 0.6% (15) | 1.5% (71) | 1.0% (41) | 1.6% (57) |  |
|  | Unknown % (n) | 16.1% (2427) | 11.2% (284) | 18.8% (896) | 18.9% (799) | 12.7% (448) |  |
|  | <missing> % (n) | 0.0% (1) | 0.0% (0) | 0.0% (1) | 0.0% (0) | 0.0% (0) |  |
| Care Home Resident | yes % (n) | 7.3% (1100) | 10.5% (267) | 5.9% (281) | 7.6% (323) | 6.5% (229) | <0.001 †† |
| Smoker | Non-smoker % (n) | 37.4% (5639) | 35.1% (894) | 40.2% (1915) | 35.7% (1513) | 37.4% (1317) | <0.001 †† |
|  | Current % (n) | 13.1% (1975) | 13.9% (353) | 10.2% (487) | 13.6% (574) | 15.9% (561) |  |
|  | Ex-smoker % (n) | 41.3% (6224) | 39.3% (1002) | 41.1% (1956) | 44.3% (1875) | 39.5% (1391) |  |
|  | Unknown % (n) | 8.1% (1223) | 11.7% (298) | 8.4% (402) | 6.4% (272) | 7.1% (251) |  |
| Heart Failure | yes % (n) | 8.8% (1324) | 13.6% (347) | 8.5% (404) | 7.8% (330) | 6.9% (243) | <0.001 †† |
| Exacerbation Of Chronic Respiratory Disease | yes % (n) | 46.3% (6978) | 53.2% (1355) | 38.8% (1848) | 46.5% (1968) | 51.3% (1807) | <0.001 †† |
| CCI Category | None (0) % (n) | 15.0% (2252) | 15.0% (381) | 16.3% (774) | 13.0% (550) | 15.5% (547) | <0.001 †† |
|  | Mild (1-2) % (n) | 27.1% (4075) | 25.3% (644) | 26.2% (1246) | 29.6% (1252) | 26.5% (933) |  |
|  | Moderate (3-4) % (n) | 17.7% (2669) | 16.7% (426) | 21.8% (1038) | 15.1% (641) | 16.0% (564) |  |
|  | Severe (5+) % (n) | 40.3% (6065) | 43.0% (1096) | 35.8% (1702) | 42.3% (1791) | 41.9% (1476) |  |
| CURB 65 Category | 0-1 (Mild) % (n) | 78.4% (11809) | 75.7% (1929) | 79.0% (3762) | 76.5% (3239) | 81.8% (2879) | <0.001 †† |
|  | 2 (Moderate) % (n) | 19.1% (2879) | 20.1% (513) | 18.3% (869) | 21.4% (908) | 16.7% (589) |  |
|  | 3-5 (Severe) % (n) | 2.5% (373) | 4.1% (105) | 2.7% (129) | 2.1% (87) | 1.5% (52) |  |
| COPD | yes % (n) | 27.8% (4190) | 33.2% (846) | 23.0% (1093) | 28.3% (1200) | 29.9% (1051) | <0.001 †† |
| Asthma | yes % (n) | 18.5% (2792) | 19.7% (501) | 15.9% (755) | 18.1% (766) | 21.9% (770) | <0.001 †† |
| Bronchiectasis | no % (n) | 95.8% (14425) | 95.3% (2428) | 96.1% (4575) | 96.1% (4068) | 95.3% (3354) | 0.14 †† |
|  | yes % (n) | 4.2% (635) | 4.7% (119) | 3.9% (184) | 3.9% (166) | 4.7% (166) |  |
|  | <missing> % (n) | 0.0% (1) | 0.0% (0) | 0.0% (1) | 0.0% (0) | 0.0% (0) |  |
| IHD | no % (n) | 87.4% (13166) | 86.2% (2195) | 89.3% (4253) | 86.8% (3673) | 86.5% (3045) | <0.001 †† |
|  | yes % (n) | 12.6% (1894) | 13.8% (352) | 10.7% (507) | 13.2% (561) | 13.5% (474) |  |
|  | <missing> % (n) | 0.0% (1) | 0.0% (0) | 0.0% (0) | 0.0% (0) | 0.0% (1) |  |
| Any Immune Compromise | yes % (n) | 13.4% (2021) | 9.9% (252) | 9.0% (429) | 15.4% (653) | 19.5% (687) | <0.001 †† |
| Diabetes Type | None % (n) | 80.8% (12172) | 79.5% (2024) | 82.5% (3929) | 80.4% (3403) | 80.0% (2816) | 0.017 †† |
|  | Type 1 % (n) | 1.4% (212) | 1.6% (41) | 1.3% (64) | 1.4% (61) | 1.3% (46) |  |
|  | Type 2 % (n) | 17.8% (2676) | 18.9% (482) | 16.1% (767) | 18.2% (770) | 18.7% (657) |  |
|  | <missing> % (n) | 0.0% (1) | 0.0% (0) | 0.0% (0) | 0.0% (0) | 0.0% (1) |  |
| CKD | None % (n) | 77.9% (11726) | 79.2% (2016) | 80.6% (3838) | 74.8% (3167) | 76.8% (2705) | <0.001 †† |
|  | Mild (CKD 1-3) % (n) | 19.1% (2881) | 17.4% (443) | 16.7% (793) | 21.9% (927) | 20.4% (718) |  |
|  | Moderate or Severe CKD (CKD 4+) % (n) | 3.0% (453) | 3.5% (88) | 2.7% (129) | 3.3% (140) | 2.7% (96) |  |
|  | <missing> % (n) | 0.0% (1) | 0.0% (0) | 0.0% (0) | 0.0% (0) | 0.0% (1) |  |
| Pneumococcal vaccination | Not received % (n) | 46.9% (7066) | 43.5% (1107) | 44.6% (2123) | 49.1% (2078) | 49.9% (1758) | <0.001 †† |
|  | PPV23 % (n) | 44.6% (6723) | 49.2% (1252) | 45.1% (2149) | 43.5% (1842) | 42.0% (1480) |  |
|  | PCV13 % (n) | 0.5% (69) | 0.0% (0) | 0.2% (9) | 1.1% (45) | 0.4% (15) |  |
|  | Unknown % (n) | 8.0% (1203) | 7.4% (188) | 10.1% (479) | 6.4% (269) | 7.6% (267) |  |
| Influenza Vaccination | Not received % (n) | 33.5% (5051) | 40.4% (1029) | 30.8% (1466) | 30.2% (1280) | 36.2% (1276) | <0.001 †† |
|  | Received % (n) | 61.1% (9197) | 52.3% (1333) | 59.6% (2837) | 67.1% (2841) | 62.1% (2186) |  |
|  | Unknown % (n) | 5.4% (807) | 7.3% (185) | 9.6% (457) | 2.7% (113) | 1.5% (52) |  |
|  | <missing> % (n) | 0.0% (6) | 0.0% (0) | 0.0% (0) | 0.0% (0) | 0.2% (6) |  |
| †, Kruskal-Wallis rank sum test (continuous); ††, Fisher's exact test (categorical)  Normal distributions determined by the Anderson-Darling test (P>0.005)  An adjusted P value of 0.0025 may be considered significant. | | | | | | |  |

In the UK, patients aged ≥65 years are eligible for Pneumococcal vaccination (PneumoVax®, PPV23) once, and annual influenza vaccine (please see Green Book23.

*Chronic kidney disease (CKD) was classified as mild if stage 1-3; moderate/severe if stage 4-5, end-stage renal failure or there was dialysis dependence.

**Influenza vaccination indicates vaccinated in the last 12 months.

aLRTD, acute lower respiratory tract disease; CCI, Charlson Comorbidity Index; CKD, chronic kidney disease; COPD, chronic obstructive pulmonary disease; IHD, ischaemic heart disease; IQR, interquartile range; PCV13, pneumococcal conjugate vaccine (13-valent); PPV23 pneumococcal polysaccharide vaccine (23-valent)

**Supplementary Data 4 (S4): Characteristics of patients hospitalised with no evidence of LRTI for the period Aug 2020 to Jul 24 by study year**

|  |  | No evidence LRTI | Year 1, 2020-21 | Year 2, 2021-22 | Year 3, 2022-23 | Year 4, 2023-24 |  |
| --- | --- | --- | --- | --- | --- | --- | --- |
| Variable | Characteristic | Value (N=7134) | Value (N=1751) | Value (N=1909) | Value (N=1677) | Value (N=1797) | P value |
| Age | Median [IQR] | 73.9 [59.9—83.5] | 75.6 [63.7—85.4] | 73 [58.5—83.1] | 73.1 [59.5—82.5] | 73.7 [59—82.8] | <0.001 † |
| Age Category | 18-34 % (n) | 6.4% (460) | 4.9% (85) | 8.1% (154) | 6.0% (100) | 6.7% (121) | <0.001 †† |
|  | 35-49 % (n) | 8.4% (599) | 6.1% (107) | 9.4% (180) | 9.1% (152) | 8.9% (160) |  |
|  | 50-64 % (n) | 17.6% (1257) | 15.8% (277) | 17.3% (330) | 19.0% (319) | 18.4% (331) |  |
|  | 65-74 % (n) | 20.4% (1458) | 21.6% (378) | 19.7% (376) | 20.6% (346) | 19.9% (358) |  |
|  | 75-84 % (n) | 25.7% (1830) | 25.5% (446) | 25.1% (479) | 26.2% (439) | 25.9% (466) |  |
|  | 85+ % (n) | 21.4% (1530) | 26.2% (458) | 20.4% (390) | 19.1% (321) | 20.1% (361) |  |
| Age Eligible for PneumoVax | 18-64 % (n) | 32.5% (2316) | 26.8% (469) | 34.8% (664) | 34.0% (571) | 34.1% (612) | <0.001 †† |
|  | 65+ % (n) | 67.5% (4818) | 73.2% (1282) | 65.2% (1245) | 66.0% (1106) | 65.9% (1185) |  |
| Gender | Female % (n) | 52.6% (3754) | 53.9% (943) | 52.0% (993) | 51.7% (867) | 52.9% (951) | 0.58 †† |
| Ethnicity | White British % (n) | 76.6% (5466) | 77.6% (1358) | 74.6% (1425) | 76.5% (1283) | 77.9% (1400) | 0.0067 †† |
|  | White other % (n) | 2.6% (189) | 2.3% (41) | 2.7% (52) | 2.4% (41) | 3.1% (55) |  |
|  | Mixed origin % (n) | 1.0% (74) | 0.7% (12) | 1.2% (23) | 1.6% (26) | 0.7% (13) |  |
|  | Black % (n) | 1.9% (136) | 1.0% (18) | 1.9% (36) | 2.5% (42) | 2.2% (40) |  |
|  | Asian % (n) | 1.7% (123) | 1.4% (25) | 2.0% (39) | 1.6% (27) | 1.8% (32) |  |
|  | Other race % (n) | 1.0% (69) | 1.1% (19) | 0.8% (16) | 0.8% (14) | 1.1% (20) |  |
|  | Unknown % (n) | 15.1% (1077) | 15.9% (278) | 16.7% (318) | 14.5% (244) | 13.2% (237) |  |
| Care Home Resident | yes % (n) | 5.0% (359) | 5.8% (101) | 4.5% (86) | 4.7% (78) | 5.2% (94) | 0.29 †† |
| Smoker | Non-smoker % (n) | 34.4% (2455) | 32.7% (573) | 35.3% (673) | 34.3% (576) | 35.2% (633) | <0.001 †† |
|  | Current % (n) | 13.0% (929) | 8.0% (140) | 12.6% (240) | 17.1% (287) | 14.6% (262) |  |
|  | Ex-smoker % (n) | 44.9% (3201) | 48.1% (843) | 46.3% (883) | 43.3% (726) | 41.7% (749) |  |
|  | Unknown % (n) | 7.7% (549) | 11.1% (195) | 5.9% (113) | 5.2% (88) | 8.5% (153) |  |
| Heart Failure | yes % (n) | 44.2% (3154) | 44.2% (774) | 43.9% (839) | 43.5% (730) | 45.1% (811) | 0.8 †† |
| Exacerbation Of Chronic Respiratory Disease | yes % (n) | 49.9% (3563) | 38.7% (677) | 51.2% (978) | 54.7% (918) | 55.1% (990) | <0.001 †† |
| CCI Category | None (0) % (n) | 12.1% (862) | 11.1% (195) | 12.2% (232) | 12.9% (216) | 12.2% (219) | <0.001 †† |
|  | Mild (1-2) % (n) | 27.8% (1982) | 27.7% (485) | 27.2% (519) | 29.2% (489) | 27.2% (489) |  |
|  | Moderate (3-4) % (n) | 10.9% (778) | 8.0% (140) | 13.6% (259) | 11.0% (184) | 10.9% (195) |  |
|  | Severe (5+) % (n) | 49.2% (3512) | 53.2% (931) | 47.1% (899) | 47.0% (788) | 49.7% (894) |  |
| CURB 65 Category | 0-1 (Mild) % (n) | 77.9% (5555) | 70.9% (1241) | 78.8% (1505) | 80.7% (1354) | 81.0% (1455) | <0.001 †† |
|  | 2 (Moderate) % (n) | 19.3% (1379) | 23.7% (415) | 18.7% (357) | 17.2% (289) | 17.7% (318) |  |
|  | 3-5 (Severe) % (n) | 2.8% (200) | 5.4% (95) | 2.5% (47) | 2.0% (34) | 1.3% (24) |  |
| COPD | yes % (n) | 31.9% (2278) | 26.6% (465) | 32.6% (622) | 35.9% (602) | 32.8% (589) | <0.001 †† |
| Asthma | yes % (n) | 19.9% (1421) | 15.5% (272) | 21.0% (400) | 20.5% (344) | 22.5% (405) | <0.001 †† |
| Bronchiectasis | yes % (n) | 3.1% (219) | 2.6% (45) | 2.6% (50) | 3.2% (54) | 3.9% (70) | 0.078 †† |
| IHD | yes % (n) | 16.5% (1174) | 18.5% (324) | 14.7% (281) | 16.3% (274) | 16.4% (295) | 0.024 †† |
| Any Immune Compromise | yes % (n) | 14.5% (1033) | 9.0% (158) | 12.4% (237) | 17.4% (291) | 19.3% (347) | <0.001 †† |
| Diabetes Type | None % (n) | 77.7% (5543) | 74.6% (1306) | 79.3% (1513) | 77.3% (1297) | 79.4% (1427) | 0.005 †† |
|  | Type 1 % (n) | 1.2% (83) | 1.0% (18) | 1.2% (23) | 1.1% (18) | 1.3% (24) |  |
|  | Type 2 % (n) | 21.1% (1508) | 24.4% (427) | 19.5% (373) | 21.6% (362) | 19.3% (346) |  |
| CKD | None % (n) | 72.0% (5136) | 69.8% (1223) | 74.3% (1419) | 71.5% (1199) | 72.1% (1295) | 0.018 †† |
|  | Mild (CKD 1-3) % (n) | 23.2% (1657) | 24.6% (431) | 21.6% (413) | 22.9% (384) | 23.9% (429) |  |
|  | Moderate or Severe CKD (CKD 4+) % (n) | 4.8% (341) | 5.5% (97) | 4.0% (77) | 5.6% (94) | 4.1% (73) |  |
| Pneumovax | Not received % (n) | 40.3% (2872) | 29.1% (510) | 38.0% (725) | 47.9% (803) | 46.4% (834) | <0.001 †† |
|  | PPV23 % (n) | 51.9% (3701) | 64.3% (1126) | 53.7% (1025) | 44.3% (743) | 44.9% (807) |  |
|  | PCV13 % (n) | 0.5% (34) | 0.0% (0) | 0.2% (4) | 1.4% (24) | 0.3% (6) |  |
|  | Unknown % (n) | 7.4% (527) | 6.6% (115) | 8.1% (155) | 6.4% (107) | 8.3% (150) |  |
| Influenza Vaccination | Not received % (n) | 27.9% (1987) | 24.6% (430) | 26.1% (498) | 28.0% (469) | 32.8% (590) | <0.001 †† |
|  | Received % (n) | 67.4% (4807) | 69.1% (1210) | 66.0% (1259) | 69.8% (1170) | 65.0% (1168) |  |
|  | Unknown % (n) | 4.7% (338) | 6.3% (111) | 8.0% (152) | 2.3% (38) | 2.1% (37) |  |
|  | <missing> % (n) | 0.0% (2) | 0.0% (0) | 0.0% (0) | 0.0% (0) | 0.1% (2) |  |
| †, Kruskal-Wallis rank sum test (continuous); ††, Fisher's exact test (categorical)  Normal distributions determined by the Anderson-Darling test (P>0.005)  An adjusted P value of 0.0025 may be considered significant. | | | | | | |  |

**Supplementary Data 5 (S5): Overall incidence of aLTRD for the period Aug 2020 to Jan 2024.**

The average incidence of aLRTD per study year is stratified by clinical presentation, causative aetiology, and age categories, and expressed as cases per 100,000 person years

|  |  |  | Year 1, 2020-21 | | Year 2, 2021-22 | | Year 3, 2022-23 | | Year 4, 2023-24 | | **4 Study Years,**  **2020-2024** | |
| --- | --- | --- | --- | --- | --- | --- | --- | --- | --- | --- | --- | --- |
|  |  | Age Group | population | incidence | population | incidence | population | incidence | population | incidence | population | incidence |
| All aLTRD | All cause | All adults | 718779 | 1228 | 729326 | 1446 | 739911 | 1341 | 751600 | 1278 | 735130 | 1324 |
|  |  | 18-64 | 577830 | 534 | 586518 | 677 | 595437 | 515 | 604913 | 541 | 591052 | 567 |
|  |  | 65+ | 140994 | 4075 | 142621 | 4612 | 144445 | 4746 | 146629 | 4318 | 143740 | 4441 |
| Pneumonia | All cause | All adults | 718725 | 673 | 728857 | 625 | 740025 | 614 | 751691 | 639 | 734800 | 638 |
|  |  | 18-64 | 577788 | 307 | 585817 | 262 | 595510 | 200 | 605062 | 249 | 590196 | 255 |
|  |  | 65+ | 140986 | 2171 | 142575 | 2120 | 144468 | 2320 | 146637 | 2246 | 143742 | 2216 |
|  | Confirmed SARS-CoV-2 | All adults | 718641 | 255 | 728332 | 262 | 739439 | 111 | 751647 | 88 | 729622 | 179 |
|  |  | 18-64 | 577888 | 170 | 585192 | 143 | 595289 | 23 | 604911 | 24 | 583795 | 90 |
|  |  | 65+ | 140931 | 602 | 142545 | 751 | 144342 | 476 | 146637 | 350 | 143174 | 545 |
|  | No evidence SARS-CoV-2 | All adults | 718776 | 418 | 729235 | 363 | 740155 | 502 | 751698 | 551 | 736801 | 459 |
|  |  | 18-64 | 577664 | 137 | 586568 | 119 | 595539 | 177 | 605078 | 225 | 593642 | 165 |
|  |  | 65+ | 141006 | 1569 | 142591 | 1368 | 144501 | 1844 | 146638 | 1896 | 143927 | 1670 |
| NP-LRTI | All cause | All adults | 718684 | 325 | 729714 | 582 | 739747 | 520 | 751500 | 426 | 735774 | 463 |
|  |  | 18-64 | 577801 | 151 | 586946 | 313 | 595379 | 230 | 604788 | 206 | 591792 | 225 |
|  |  | 65+ | 140973 | 1036 | 142672 | 1687 | 144405 | 1715 | 146619 | 1331 | 143819 | 1443 |
|  | Confirmed SARS-CoV-2 | All adults | 718756 | 44 | 730405 | 312 | 739217 | 194 | 751639 | 139 | 736554 | 172 |
|  |  | 18-64 | 578300 | 20 | 587368 | 185 | 595080 | 68 | 604934 | 52 | 591320 | 81 |
|  |  | 65+ | 140937 | 138 | 142804 | 835 | 144292 | 714 | 146636 | 496 | 144063 | 546 |
|  | No evidence SARS-CoV-2 | All adults | 718673 | 281 | 728914 | 270 | 740063 | 326 | 751433 | 287 | 735312 | 291 |
|  |  | 18-64 | 577723 | 131 | 586337 | 128 | 595504 | 162 | 604739 | 154 | 592059 | 144 |
|  |  | 65+ | 140979 | 897 | 142542 | 852 | 144485 | 1001 | 146608 | 835 | 143670 | 897 |
| No evidence LRTI | All cause | All adults | 719069 | 231 | 729605 | 239 | 739986 | 207 | 751528 | 213 | 734733 | 223 |
|  |  | 18-64 | 578062 | 75 | 587003 | 102 | 595423 | 84 | 604778 | 85 | 591639 | 87 |
|  |  | 65+ | 141042 | 869 | 142638 | 805 | 144468 | 711 | 146624 | 741 | 143587 | 782 |

aLRTD, acute lower respiratory tract disease; LRTI, lower respiratory tract infection; NP-LRTI, non-pneumonic lower respiratory tract infection

**Supplementary Data 6 (S6): Outcomes of aLTRD in different clinical presentations for the period Aug 2020 to Jul 2021**

|  |  | All aLRTD | Pneumonia | NP-LRTI | No evidence LRTI |  |
| --- | --- | --- | --- | --- | --- | --- |
| Variable | Characteristic | Value (N=42834) | Value (N=20639) | Value (N=15061) | Value (N=7134) | P value |
| Length Of Stay (days) | Median [IQR] | 5 [2—11] | 6 [3—13] | 3 [1—9] | 4 [1—9] | <0.001 † |
| ICU Admission | confirmed % (n) | 2.6% (1106) | 4.3% (897) | 1.0% (148) | 0.9% (61) | <0.001 †† |
| ICU Duration (days) | Median [IQR] | 7 [3—12.5] | 7.5 [4—14] | 4 [2—7] | 4 [2—9] | — ††† |
| Death Within 30 Days | confirmed % (n) | 9.8% (4198) | 14.7% (3039) | 4.3% (653) | 7.1% (506) | <0.001 †† |
| Death Within 1 Year | confirmed % (n) | 25.9% (11081) | 32.1% (6628) | 18.0% (2713) | 24.4% (1740) | <0.001 †† |
| †, Kruskal-Wallis rank sum test (continuous); ††, Fisher's exact test (categorical); †††, Not calculated due to missing values (continuous) Normal distributions determined by the Anderson-Darling test (P>0.005) An adjusted P value of 0.01 may be considered significant. | | | | | |  |

aLRTD, acute lower respiratory tract disease; ICU, intensive care unit; IQR, interquartile range; LRTI, lower respiratory tract infection; NP-LRTI, non-pneumonic lower respiratory tract infection

*Only individuals with complete follow-up (1 year for mortality within 1 year; 30 days for mortality within 30 days) were included.

The overall outcomes of aLRTD with different clinical presentations for the period August 2020 to July 2024 are summarized in Table 3. The median length of hospital stay for all aLRTD cases was 5 days, with pneumonia patients experiencing the longest stays at 6 days, followed by those with no evidence of LRTI at 4 days, and NP-LRTI at 3 days. ICU admission rates were highest for pneumonia cases at 4.3%, compared to 1.0% for NP-LRTI and 0.9% for cases with no evidence of LRTI, with the median ICU duration being 7 days for all aLRTD cases, with pneumonia cases having the longest stays. Overall, mortality rates within 30 days were highest for pneumonia at 14.7%, followed by cases with no evidence of LRTI at 7.1%, and NP-LRTI at 4.3%, with an overall 30-day mortality rate of 9.8%. The 1-year mortality rate was also highest for pneumonia (32.1%), with an overall rate for all aLRTD of 25.9%. Detailed results for each study year can be found in Supplementary Data S7-S10.

**Supplementary Data 7 (S7): Outcomes of aLTRD in different clinical presentations for the period Aug 2020 to Jul 2021**

|  |  | All aLRTD | Pneumonia | NP-LRTI | No evidence LRTI |  |
| --- | --- | --- | --- | --- | --- | --- |
| Variable | Characteristic | Value (N=9585) | Value (N=5285) | Value (N=2546) | Value (N=1754) | P value |
| Length Of Stay | Median [IQR] | 5 [2—11] | 6 [3—12] | 3 [1—8] | 4 [1—10] | <0.001 † |
| ICU Admission | confirmed % (n) | 3.2% (306) | 5.0% (264) | 0.9% (23) | 1.1% (19) | <0.001 †† |
| ICU Duration | Median [IQR] | 7 [4—13] | 7 [4—14] | 3 [1.5—7] | 5 [3.5—8] | — ††† |
| Death Within 30 Days | confirmed % (n) | 11.4% (1092) | 15.5% (818) | 5.7% (145) | 7.4% (129) | <0.001 †† |
| Death Within 1 Year | confirmed % (n) | 27.4% (2627) | 30.9% (1635) | 21.8% (554) | 25.0% (438) | <0.001 †† |
| †, Kruskal-Wallis rank sum test (continuous); ††, Fisher's exact test (categorical); †††, Not calculated due to missing values (continuous)  Normal distributions determined by the Anderson-Darling test (P>0.005)  An adjusted P value of 0.01 may be considered significant. | | | | | |  |

aLRTD, acute lower respiratory tract disease; ICU, intensive care unit; IQR, interquartile range; LRTI, lower respiratory tract infection; NP-LRTI, non-pneumonic lower respiratory tract infection

*Only individuals with complete follow-up (1 year for mortality within 1 year; 30 days for mortality within 30 days) were included.

**Supplementary Data 8 (S8): Outcomes of aLTRD in different clinical presentations for the period Aug 2021 to Jul 2022**

|  |  | All aLRTD | Pneumonia | NP-LRTI | No evidence LRTI |  |
| --- | --- | --- | --- | --- | --- | --- |
| Variable | Characteristic | Value (N=11770) | Value (N=5103) | Value (N=4765) | Value (N=1902) | P value |
| Length Of Stay | Median [IQR] | 5 [2—12] | 7 [3—14] | 4 [1—10] | 4 [1—9] | <0.001 † |
| ICU Admission | confirmed % (n) | 3.2% (372) | 5.5% (281) | 1.4% (68) | 1.2% (23) | <0.001 †† |
| ICU Duration | Median [IQR] | 7 [3—14] | 9 [4—16] | 4 [2—7.25] | 4 [3—10] | — ††† |
| Death Within 30 Days | confirmed % (n) | 9.8% (1149) | 15.5% (792) | 4.5% (215) | 7.5% (142) | <0.001 †† |
| Death Within 1 Year | confirmed % (n) | 26.4% (3110) | 33.7% (1718) | 18.4% (876) | 27.1% (516) | <0.001 †† |
| †, Kruskal-Wallis rank sum test (continuous); ††, Fisher's exact test (categorical); †††, Not calculated due to missing values (continuous)  Normal distributions determined by the Anderson-Darling test (P>0.005)  An adjusted P value of 0.01 may be considered significant. | | | | | |  |

aLRTD, acute lower respiratory tract disease; ICU, intensive care unit; IQR, interquartile range; LRTI, lower respiratory tract infection; NP-LRTI, non-pneumonic lower respiratory tract infection

*Only individuals with complete follow-up (1 year for mortality within 1 year; 30 days for mortality within 30 days) were included.

**Supplementary Data 9 (S9): Outcomes of aLTRD in different clinical presentations for the period Aug 2022 to Jul 2023**

|  |  | All aLRTD | Pneumonia | NP-LRTI | No evidence LRTI |  |
| --- | --- | --- | --- | --- | --- | --- |
| Variable | Characteristic | Value (N=11019) | Value (N=5047) | Value (N=4269) | Value (N=1703) | P value |
| Length Of Stay | Median [IQR] | 5 [2—12] | 6 [3—14] | 4 [1—11] | 4 [1—9] | <0.001 † |
| ICU Admission | confirmed % (n) | 2.2% (244) | 3.9% (196) | 0.9% (37) | 0.6% (11) | <0.001 †† |
| ICU Duration | Median [IQR] | 6 [3—10] | 6 [3—12] | 3 [2—7] | 3 [2—4] | — ††† |
| Death Within 30 Days | confirmed % (n) | 9.1% (1005) | 14.8% (746) | 3.8% (163) | 5.6% (96) | <0.001 †† |
| Death Within 1 Year | confirmed % (n) | 27.5% (3033) | 35.5% (1794) | 19.4% (829) | 24.1% (410) | <0.001 †† |
| †, Kruskal-Wallis rank sum test (continuous); ††, Fisher's exact test (categorical); †††, Not calculated due to missing values (continuous)  Normal distributions determined by the Anderson-Darling test (P>0.005)  An adjusted P value of 0.01 may be considered significant. | | | | | |  |

aLRTD, acute lower respiratory tract disease; ICU, intensive care unit; IQR, interquartile range; LRTI, lower respiratory tract infection; NP-LRTI, non-pneumonic lower respiratory tract infection

*Only individuals with complete follow-up (1 year for mortality within 1 year; 30 days for mortality within 30 days) were included.

**Supplementary Data 10 (S10): Outcomes of aLTRD in different clinical presentations for the period Aug 2023 to Jul 2024**

|  |  | All aLRTD | Pneumonia | NP-LRTI | No evidence LRTI |  |
| --- | --- | --- | --- | --- | --- | --- |
| Variable | Characteristic | Value (N=10434) | Value (N=5192) | Value (N=3471) | Value (N=1771) | P value |
| Length Of Stay | Median [IQR] | 5 [1—11] | 6 [2—13] | 3 [1—8] | 4 [1—9] | <0.001 † |
| ICU Admission | confirmed % (n) | 1.7% (182) | 3.0% (154) | 0.6% (20) | 0.5% (8) | <0.001 †† |
| ICU Duration | Median [IQR] | 7 [3—12] | 7 [4—12] | 3.5 [2—6.25] | 7 [2—12] | — ††† |
| Death Within 30 Days | confirmed % (n) | 9.1% (950) | 13.2% (684) | 3.7% (128) | 7.8% (138) | <0.001 †† |
| Death Within 1 Year | confirmed % (n) | 22.1% (2303) | 28.5% (1481) | 12.9% (449) | 21.1% (373) | <0.001 †† |
| †, Kruskal-Wallis rank sum test (continuous); ††, Fisher's exact test (categorical); †††, Not calculated due to missing values (continuous)  Normal distributions determined by the Anderson-Darling test (P>0.005)  An adjusted P value of 0.01 may be considered significant. | | | | | |  |

aLRTD, acute lower respiratory tract disease; ICU, intensive care unit; IQR, interquartile range; LRTI, lower respiratory tract infection; NP-LRTI, non-pneumonic lower respiratory tract infection

*Only individuals with complete follow-up (1 year for mortality within 1 year; 30 days for mortality within 30 days) were included.

**Supplementary Data 11 (S11): Kaplan-Meier unadjusted survival curves**

*Cases are stratified by clinical presentation (pneumonia, red; NP-LRTI, green; no evidence LRTI, blue) for (A) death and (B) discharge within 30 days of hospitalisation. Censored at the earliest of 30 days or 31st July 2024 ; and discharge, censored at 30 days or death.*

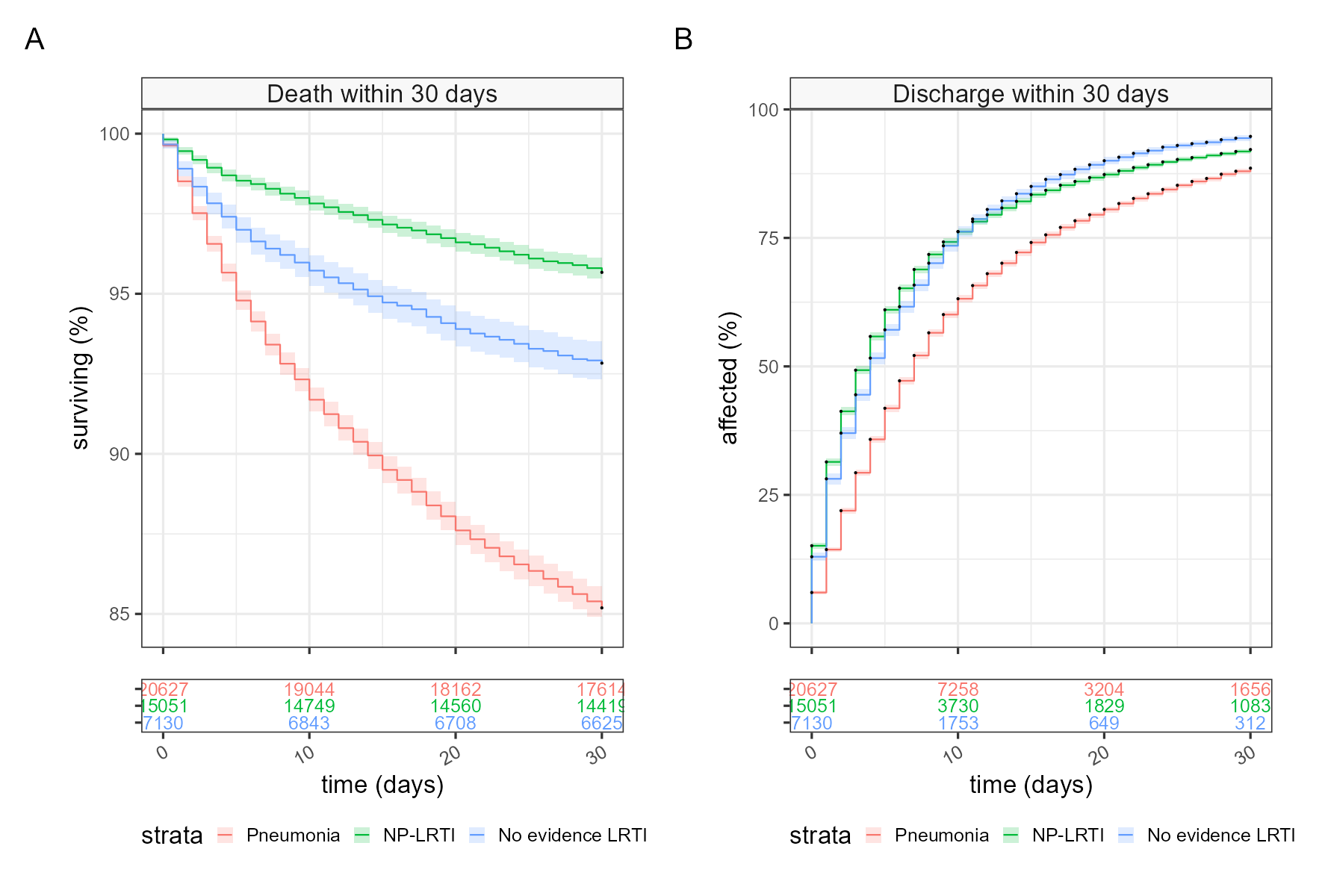
